# A Meta-Review about Medical 3D Printing

**DOI:** 10.1101/2024.04.11.23300674

**Authors:** Melissa Meister, Gijs Luijten, Christina Gsaxner, Kunpeng Xie, Lennart J. Gruber, Jianning Li, Antonio Pepe, Yao Li, Ashkan Rashad, Constantin Seibold, Fin H. Bahnsen, Moon Kim, Nino Fijačko, Frank Hölzle, Malik Sallam, Rainer Röhrig, Gregor Štiglic, Julius Keyl, Jens Kleesiek, Victor Alves, Xiaojun Chen, Behrus Puladi, Jan Egger

## Abstract

In recent years, 3D printing (3DP) has gained importance in various fields. This technology has numerous applications, particularly in medicine. This contribution provides an overview on the state of the art of 3DP in medicine and showcases its current use in different medical disciplines and for medical education. In this meta-review, we provide a detailed listing of systematic reviews on this topic as this technology has become increasingly applied in modern medicine. We identified 134 relevant systematic reviews on medical 3DP in the medical search engine PubMed until 2023. 3DP has applications in various medical specialties, but is mainly used in orthopedics, oral and maxillofacial surgery, dentistry, cardiology and neurosurgery. In surgical contexts, the adoption of 3DP contributes to a reduction in operation time, reduced blood loss, minimized fluoroscopy time and an overall improved surgical outcome. Nevertheless, the primary use of 3DP is observed in non-invasive applications, particularly in the creation of patient-specific models (PSM). These PSMs enhance the visualization of patients’ anatomy and pathology, thereby facilitating surgical planning and execution, medical education and patient counseling. The current significance of 3DP in medicine offers a compelling perspective on the potential for more individualized and personalized medical treatments in the future.

## 1 Introduction

**3**D-printing (3DP) has numerous applications in medical settings. This contribution offers an overview of 3DP in medicine and showcases its current use in medical disciplines such as oral and maxillofacial surgery or medical education. We provide a comprehensive compilation of systematic reviews on this topic, emphasizing the integration of this technology into modern medicine. We conducted a search for the keyword “3D printing” in the medical search engine PubMed, which delivered almost 30,000 articles. The substantial volume of existing literature underscores the significance of this topic. Consequently, we narrowed our focus to systematic reviews on medical 3DP, resulting in 204 publications in the last search conducted at the beginning of 2024. Following the exclusion of non-relevant publications, we provide a meta-review of a final set of 134 systematic reviews. From each systematic review, we provide information about the targeted medical field, organ, pathology and/or procedure, the used printing materials and the associated costs in terms of financial investment and time. We further report the number of included publications on the reviews, the used search keywords and conclusions, as well as the quality of the systematic review.

## 2 Reviews IN Medical 3D Printing

### 2.1 Search Strategy

An electronic search was performed in the database PubMed, with the search term “3D printing” filtered for systematic reviews at January 28th, 2024. Inclusion criteria were systematic reviews, with or without meta-analysis, written in English, focusing on 3D printing in clinical use or medical education, or comparing 3DP with other approaches. Exclusion criteria were publications on bioprinting only, animal studies only, focus on material properties or printing technology, 3DP as one of many topics, and no English full text. The literature search was performed independently by M.L. and B.P. and was resolved by G.L. or J.E. in case of disagreement. Rayyan.ai was used as a tool. The initial database search yielded 261 papers from 2015 to 2024, which were then screened based on their title and abstract. Out of these, 114 records were excluded because they did not meet the criteria. Of 146 papers, 13 further papers were excluded, 1 was not accessible, 2 were the wrong publication type (i.e. not a systematic review), 6 did not focus on 3DP, 3 were material studies and 1 was an animal study, leaving 134 papers in our meta-review (Figure 1).

**Figure 1.**
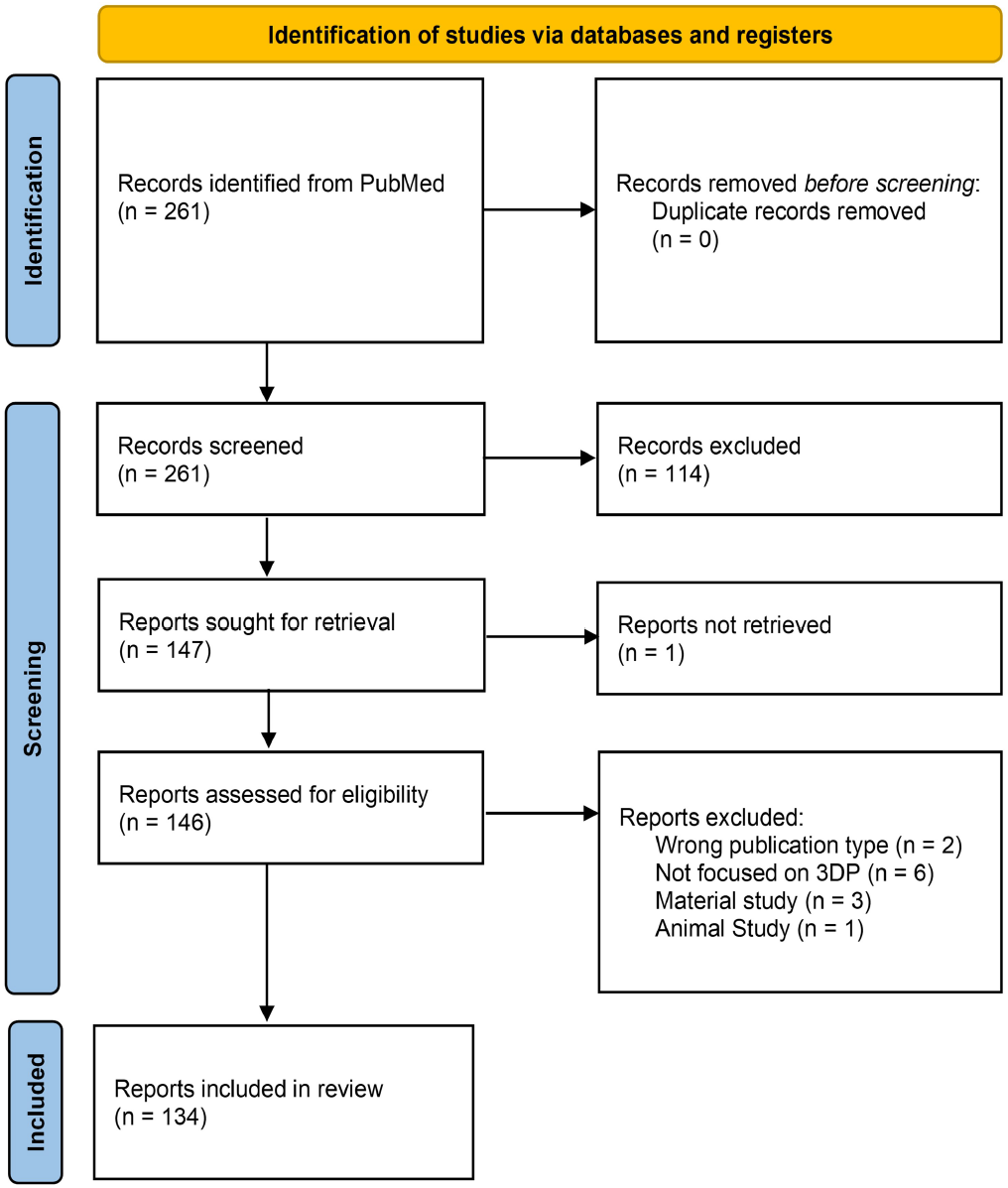
PRISMA flowchart of the screening process.

### 2.2 Data extraction

We collected the full-text articles in Mendeley. Data extraction from the 134 papers was performed by M.M. and G.L. In case of discrepancies, B.P. and J.E. were consolidated and then classified according to the degree of invasiveness (Figure 2): non-invasive (see Table 1), on-body (see Table 2), invasive non-permanent (see Table 3), and invasive permanent devices (see Table 4). (Note: The classification does not necessarily reflect the Medical Device Regulation, e.g. Class I, IIa, IIb, III). Where papers could be classified in more than one category, they were listed in several tables (n= 8). However, eight publications could not be classified in any category, despite their clinical relevance. These were publications that focused primarily on printing materials in the clinical setting. The following information was extracted from each review and collected in the tables from left to right: author, medical field, organ, pathology described (Path.), medical procedure, material used for printing, application (preoperative planning (P), teaching (T) or patient counseling (C)), authors’ region, the costs associated with hardware or the printing process, the time saved by using the technology or the time of the whole printing process and the conclusion of the review. We further include information on whether the study is a meta-analysis (MA), (‘yes’ if yes, ‘no’ if not), the number of reviewed papers included (IP), the keywords from the paper (KW). Finally, we assess the quality of the study: whether it followed the PRISMA guidelines, whether the review was registered (e.g. in PROSPERO) and whether it included a bias assessment (BA).

**Table 1:**
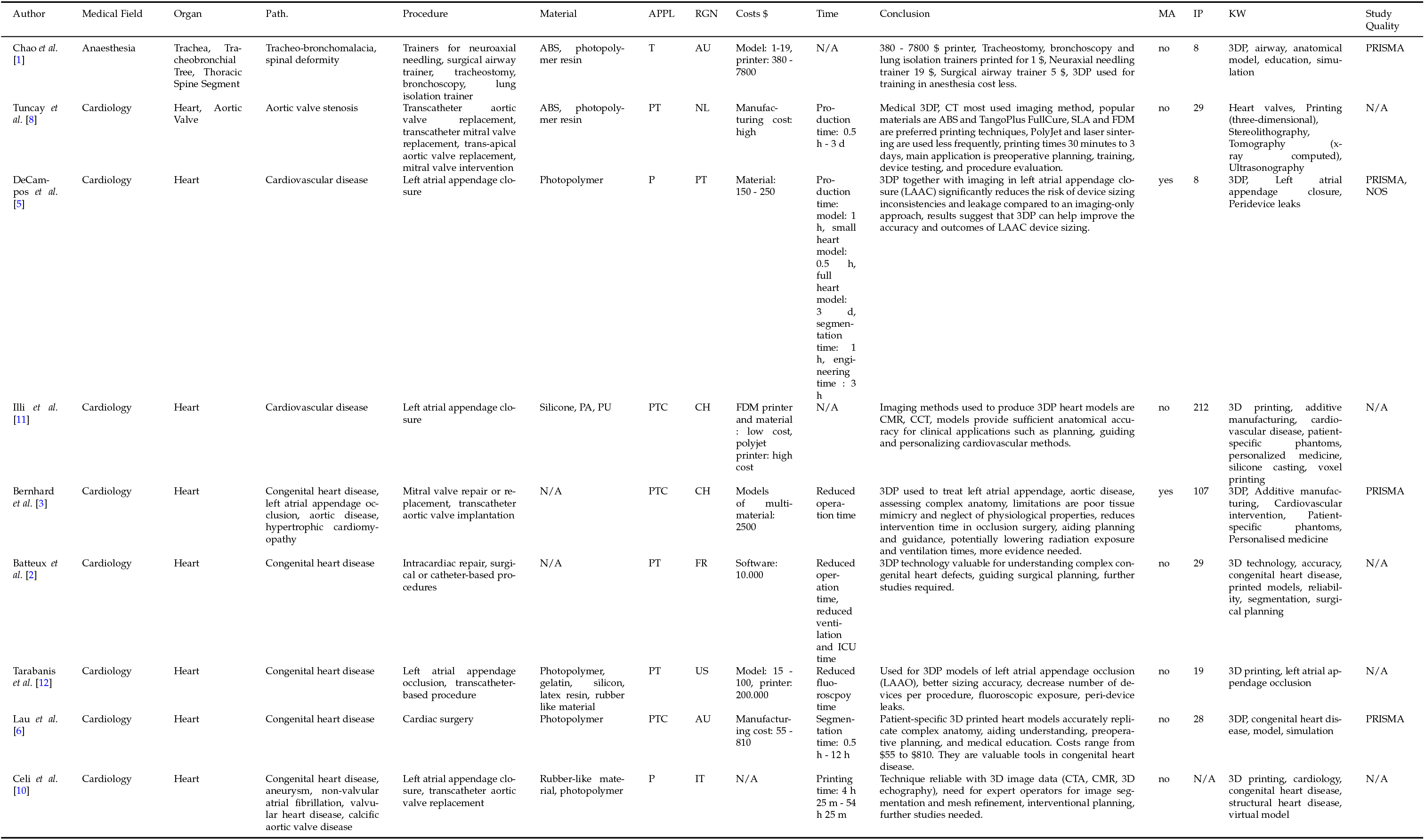
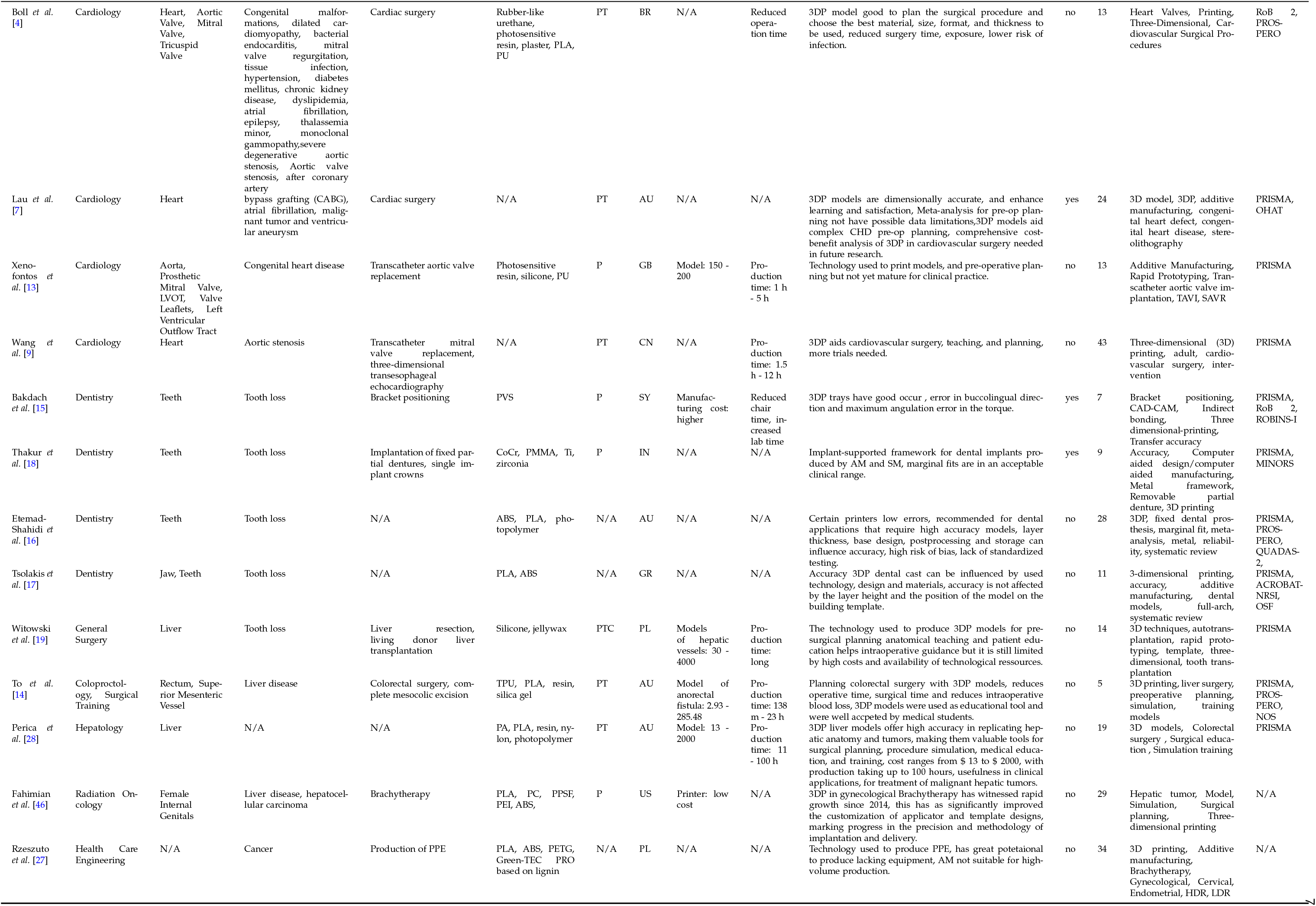
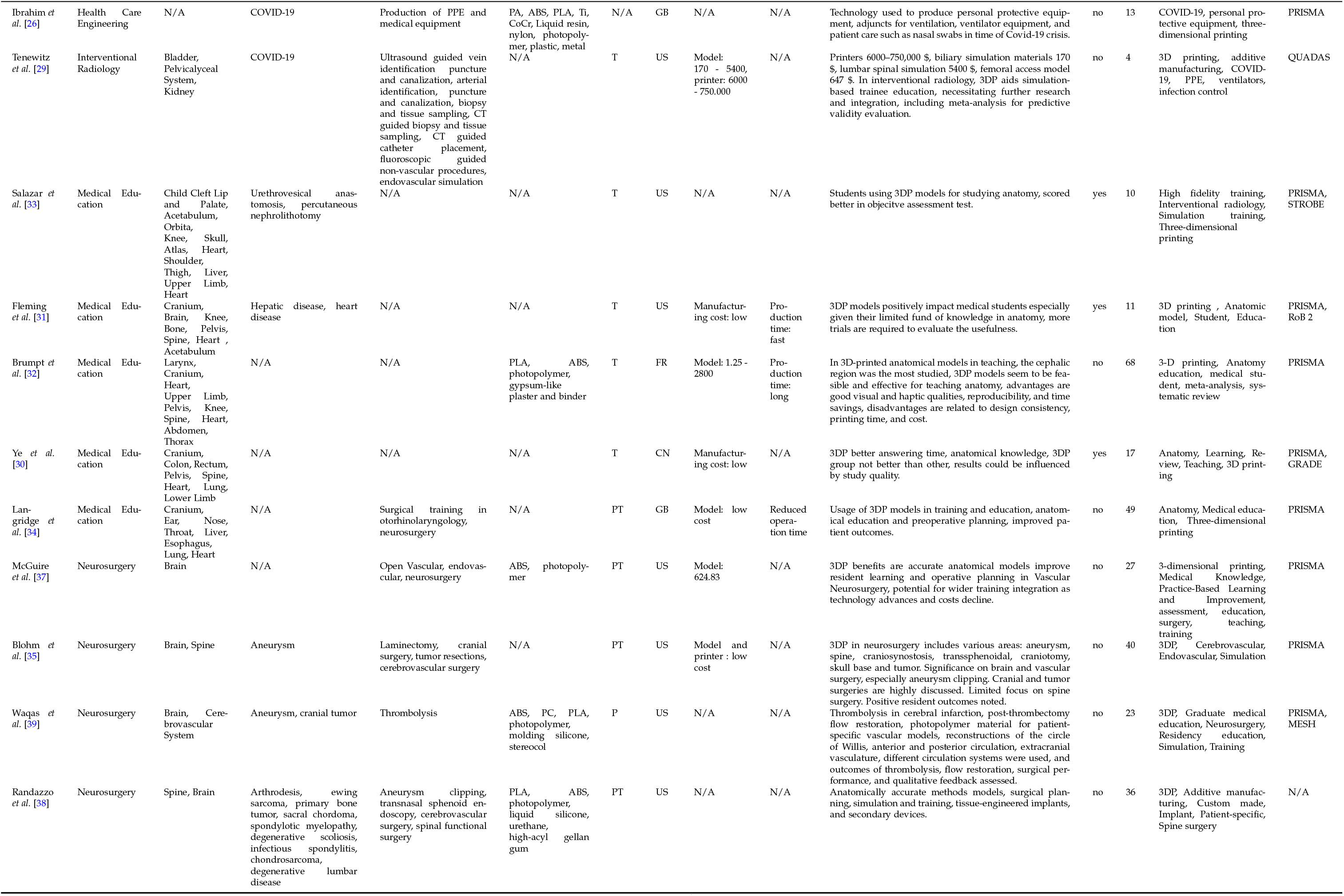
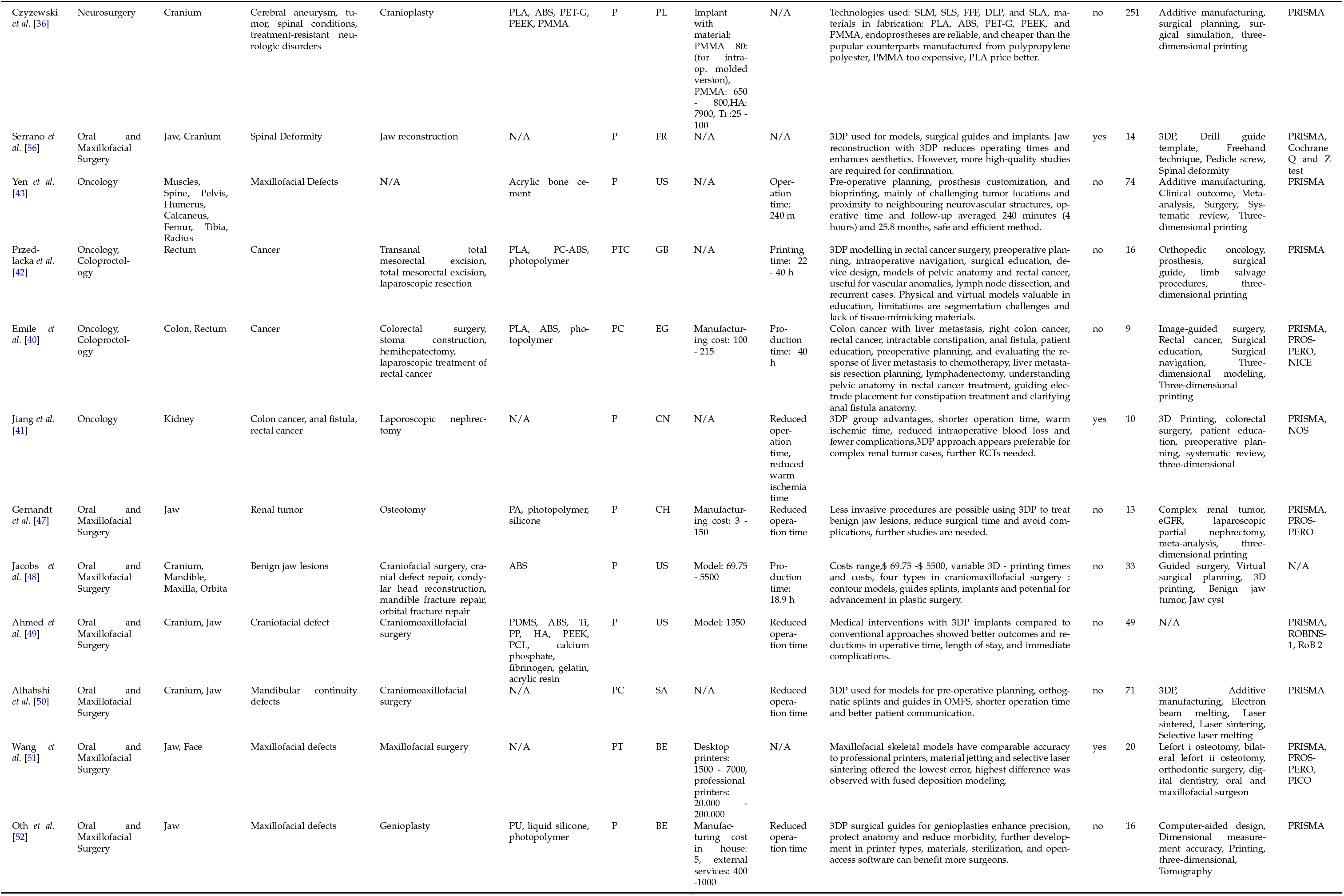
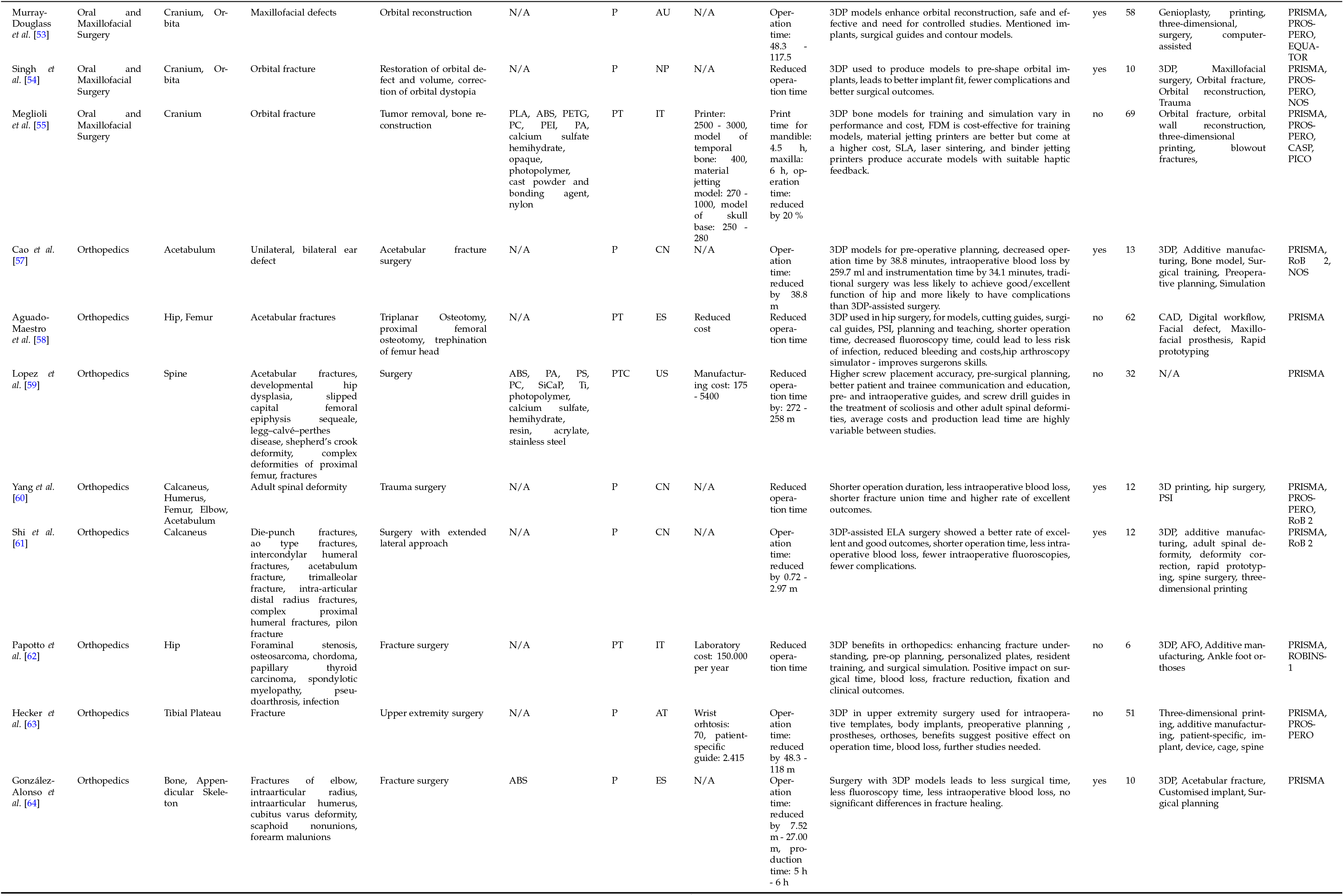
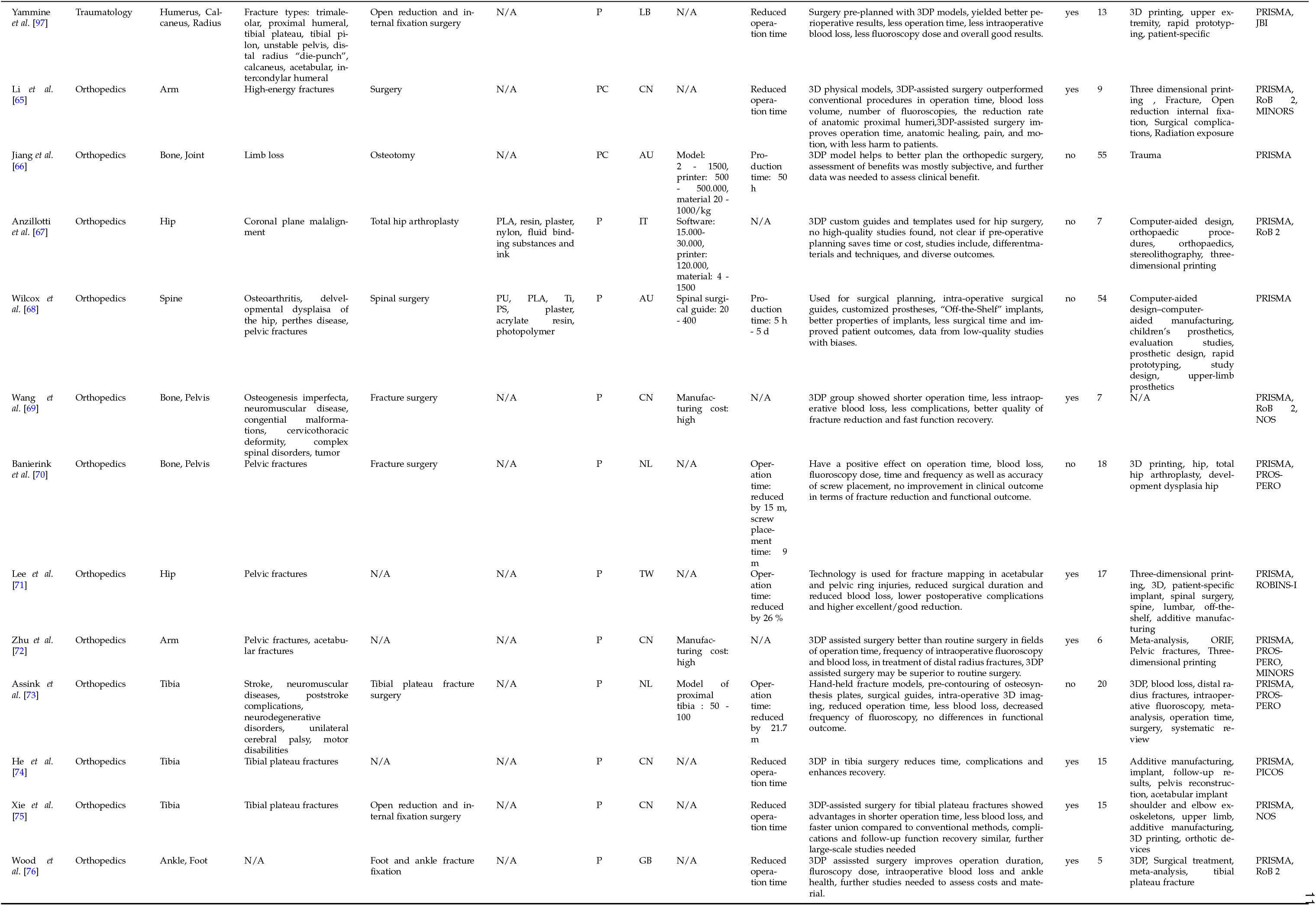
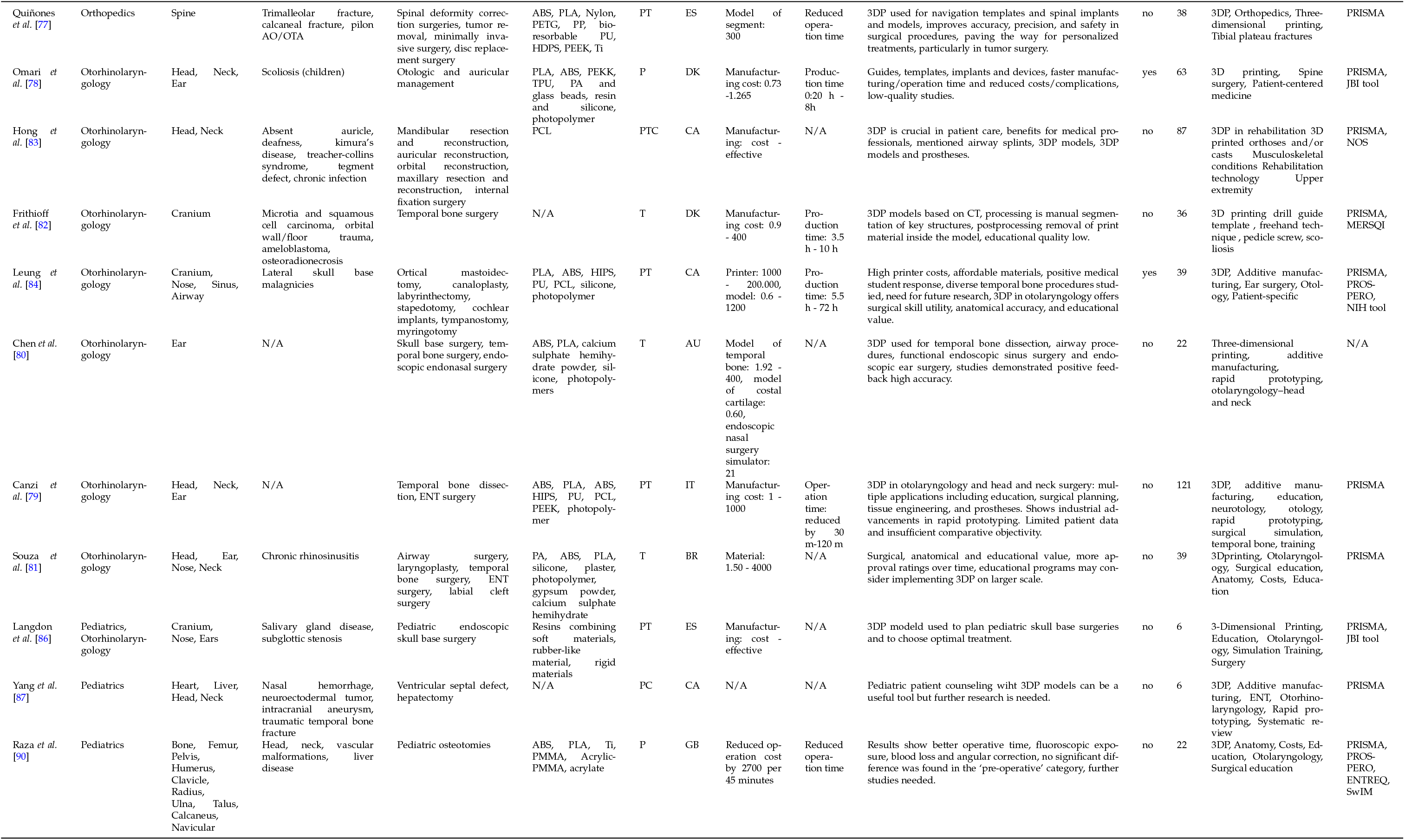
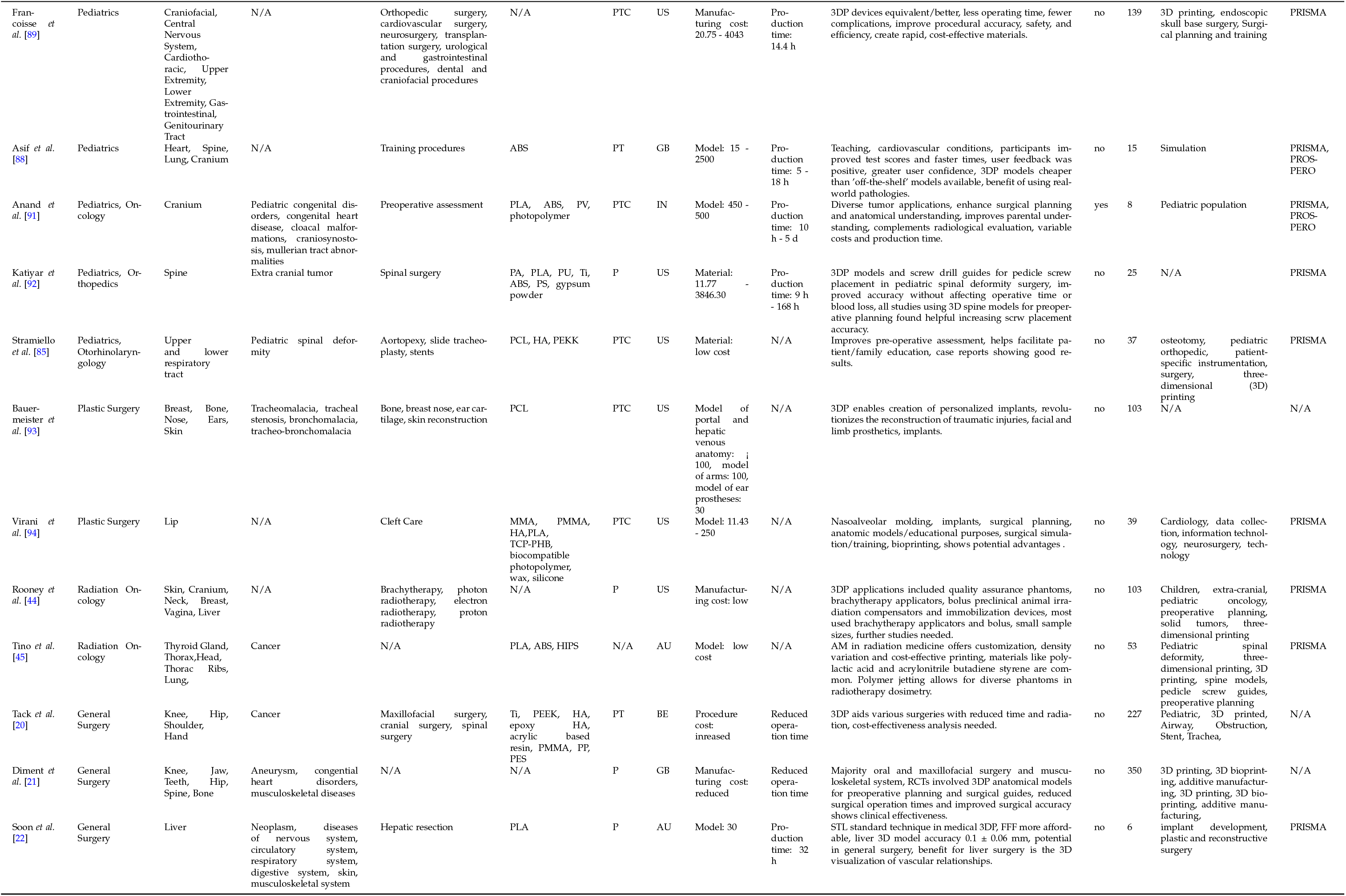
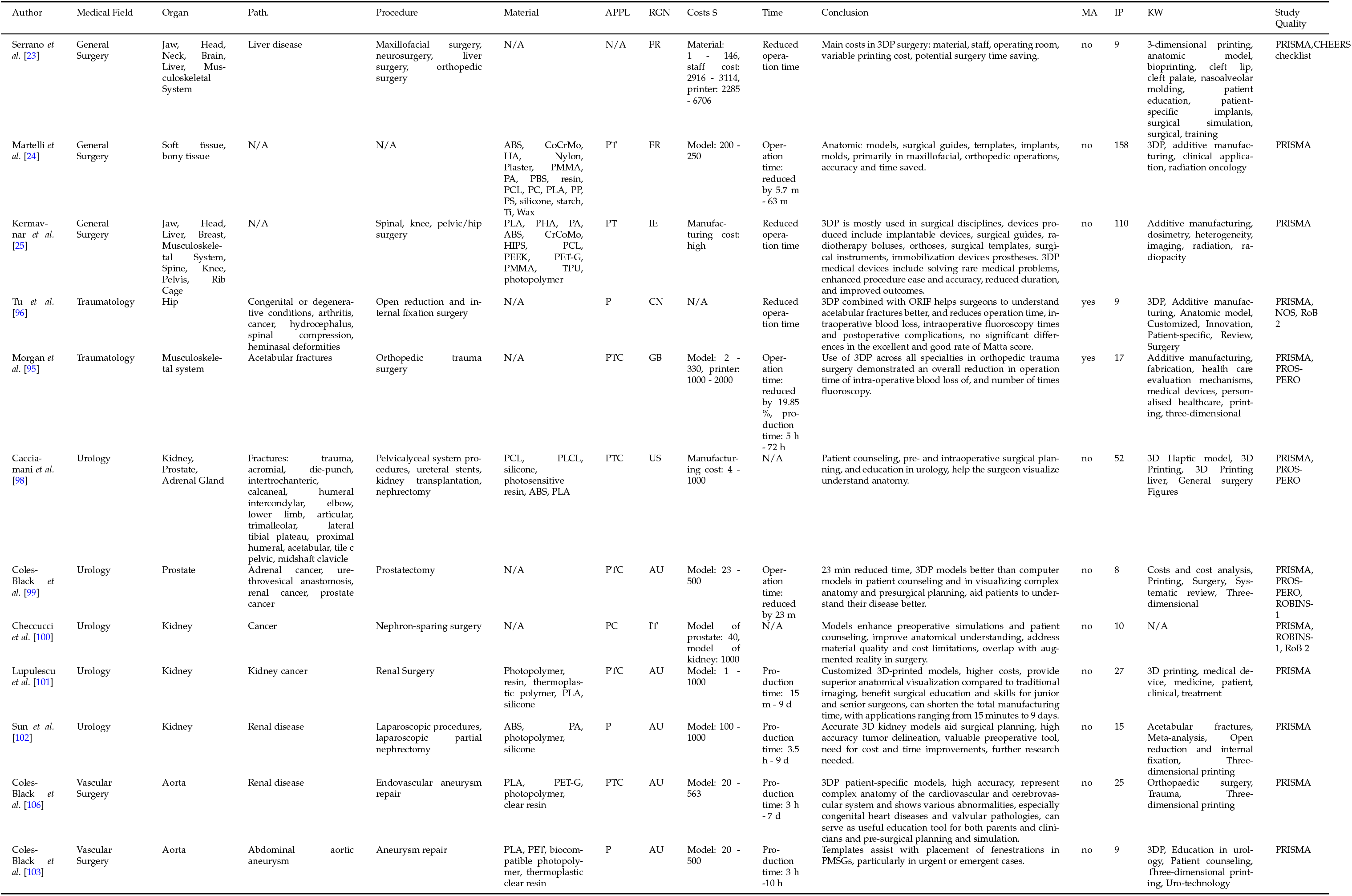
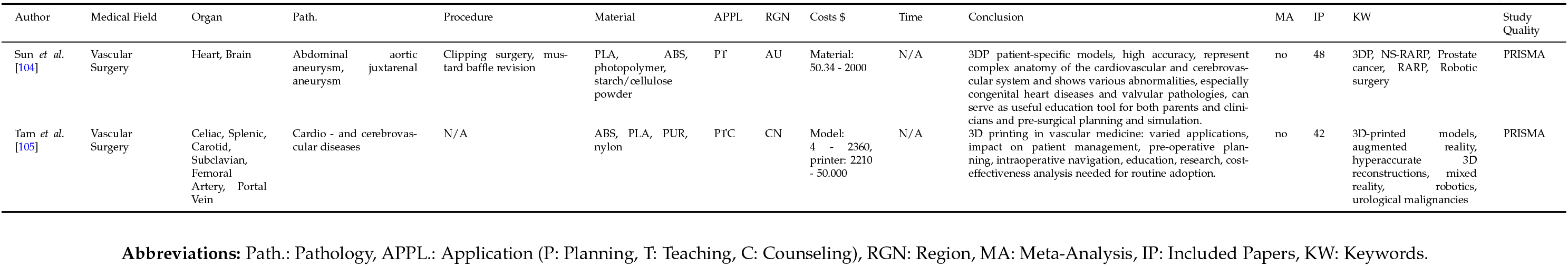
Systematic Reviews of Non-Invasive Applications.

**Table 2:**
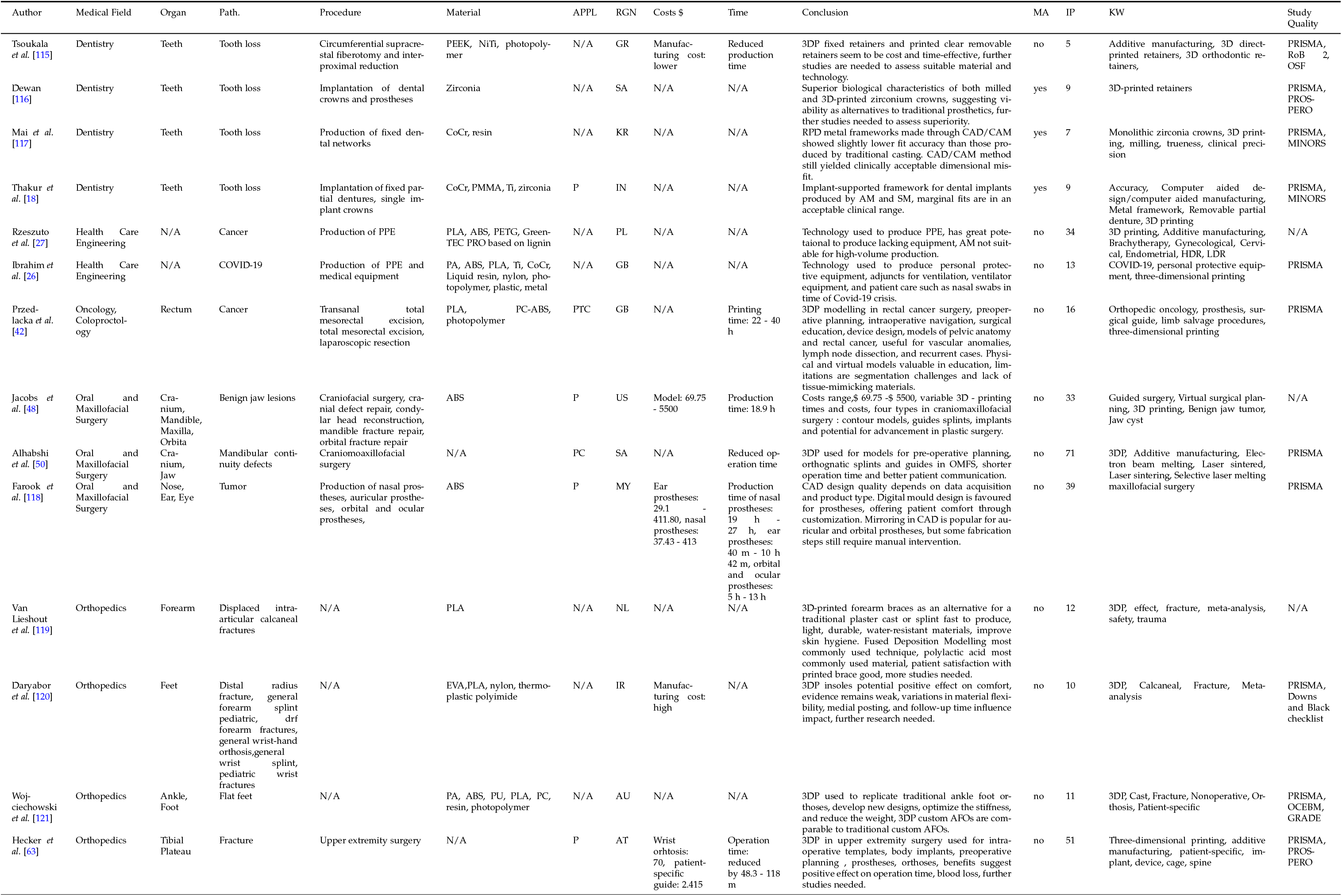
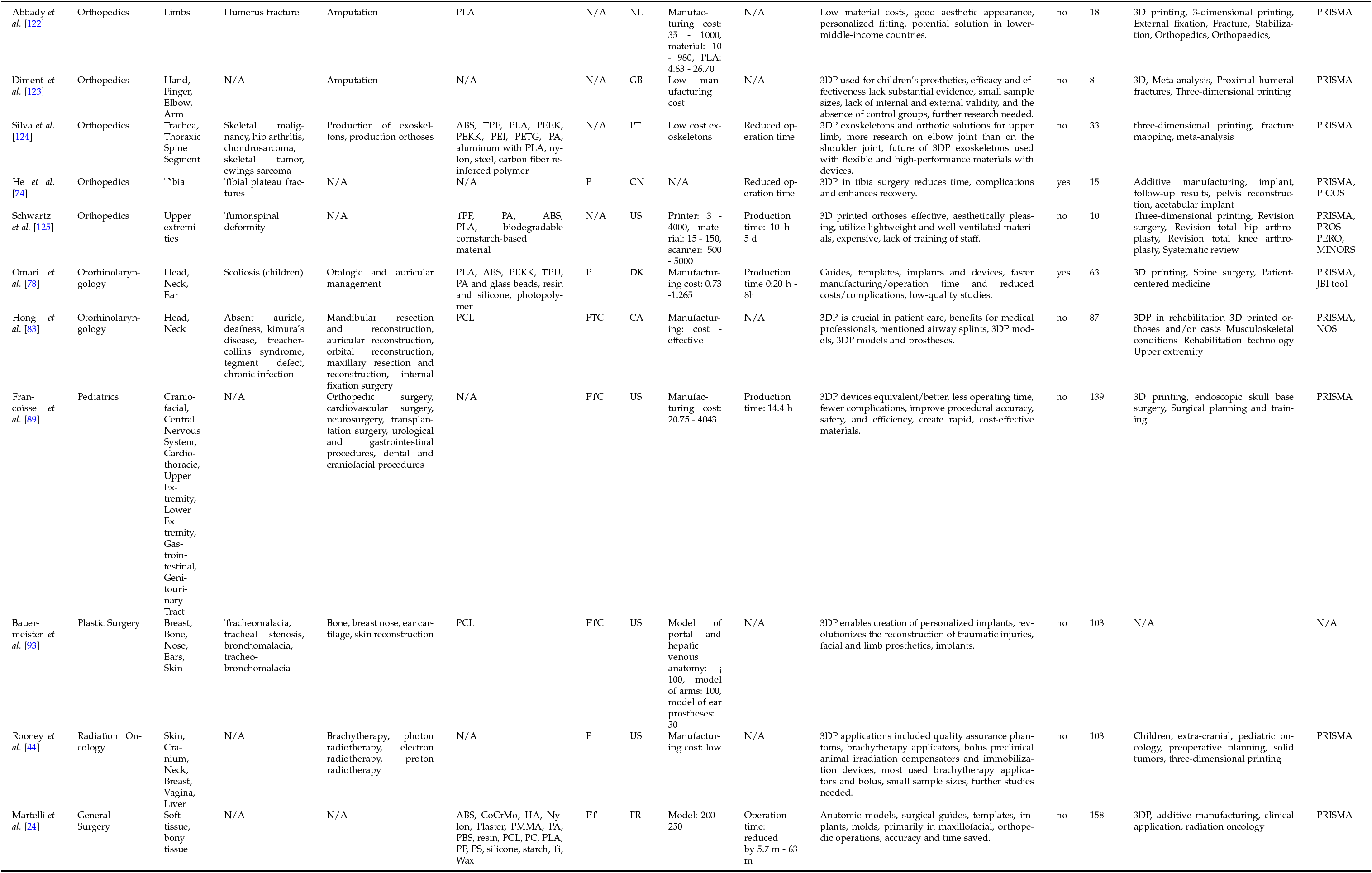
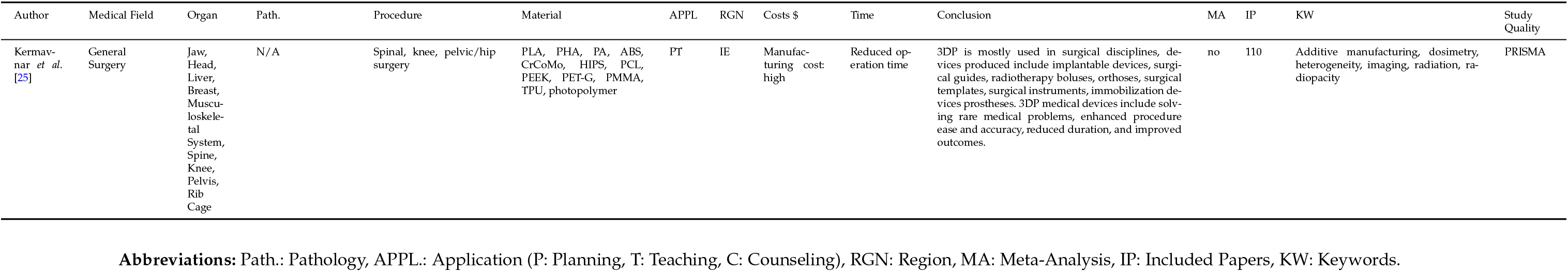
Systematic Reviews of On-Body Applications.

**Table 3:**
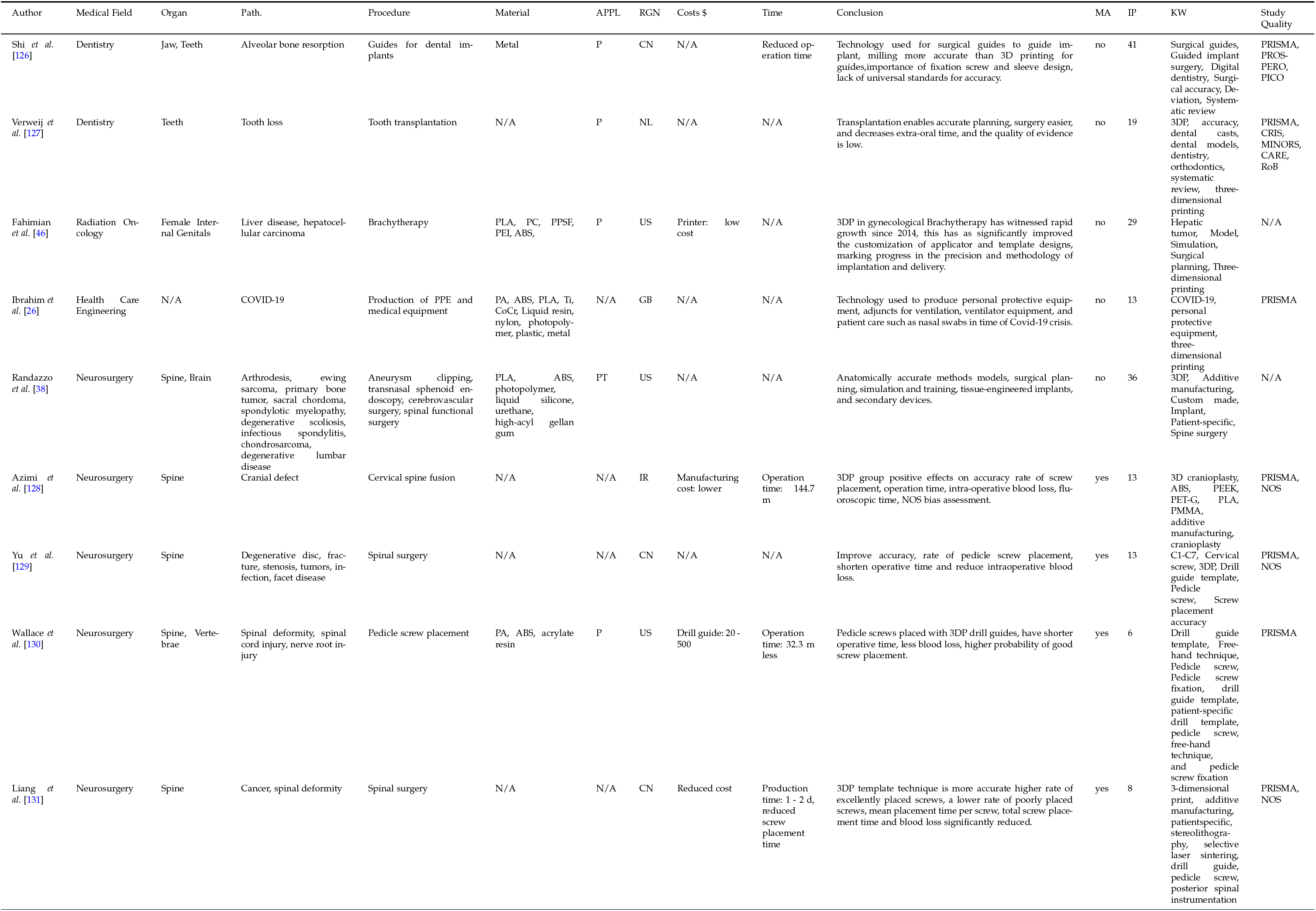
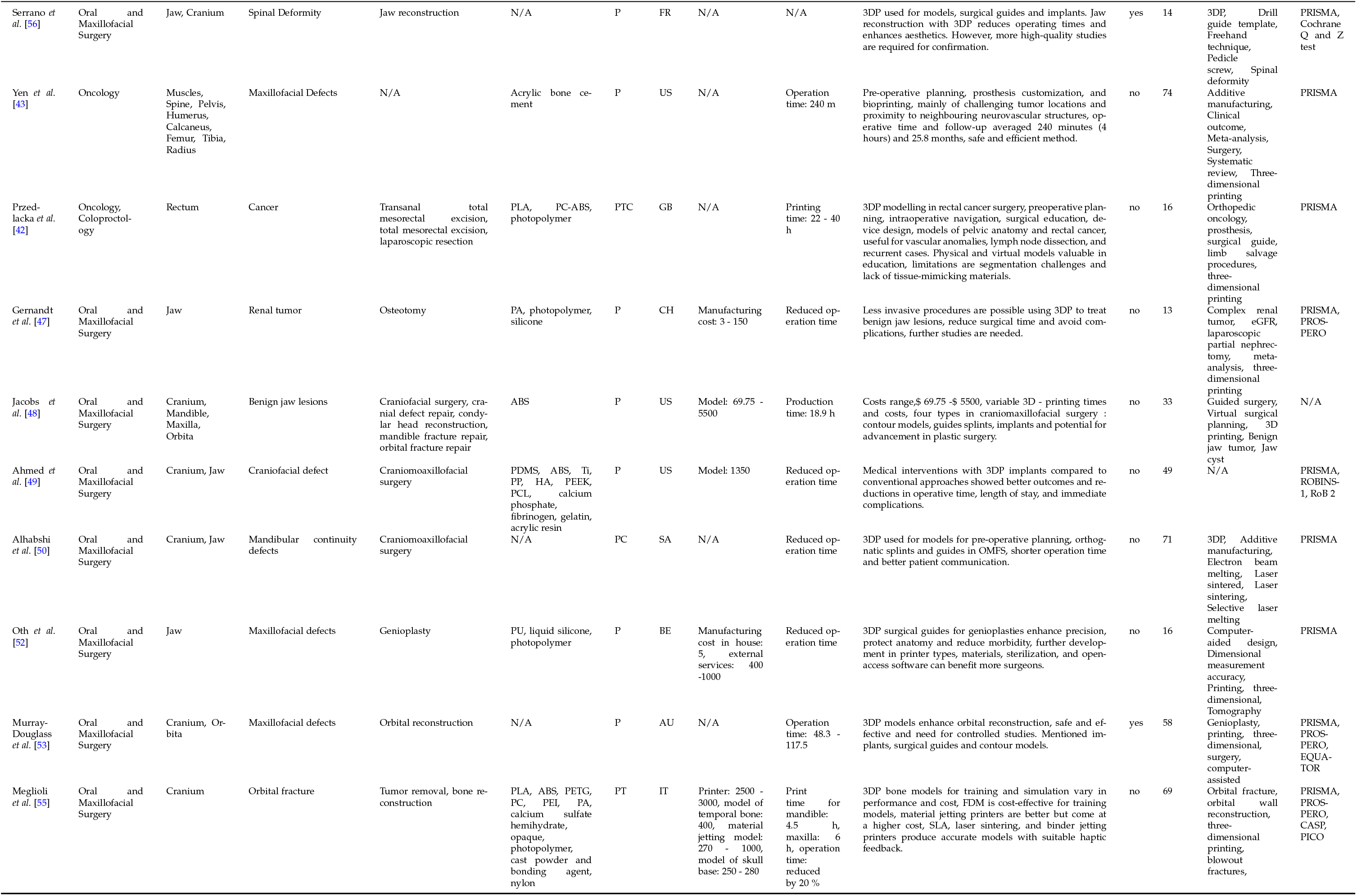
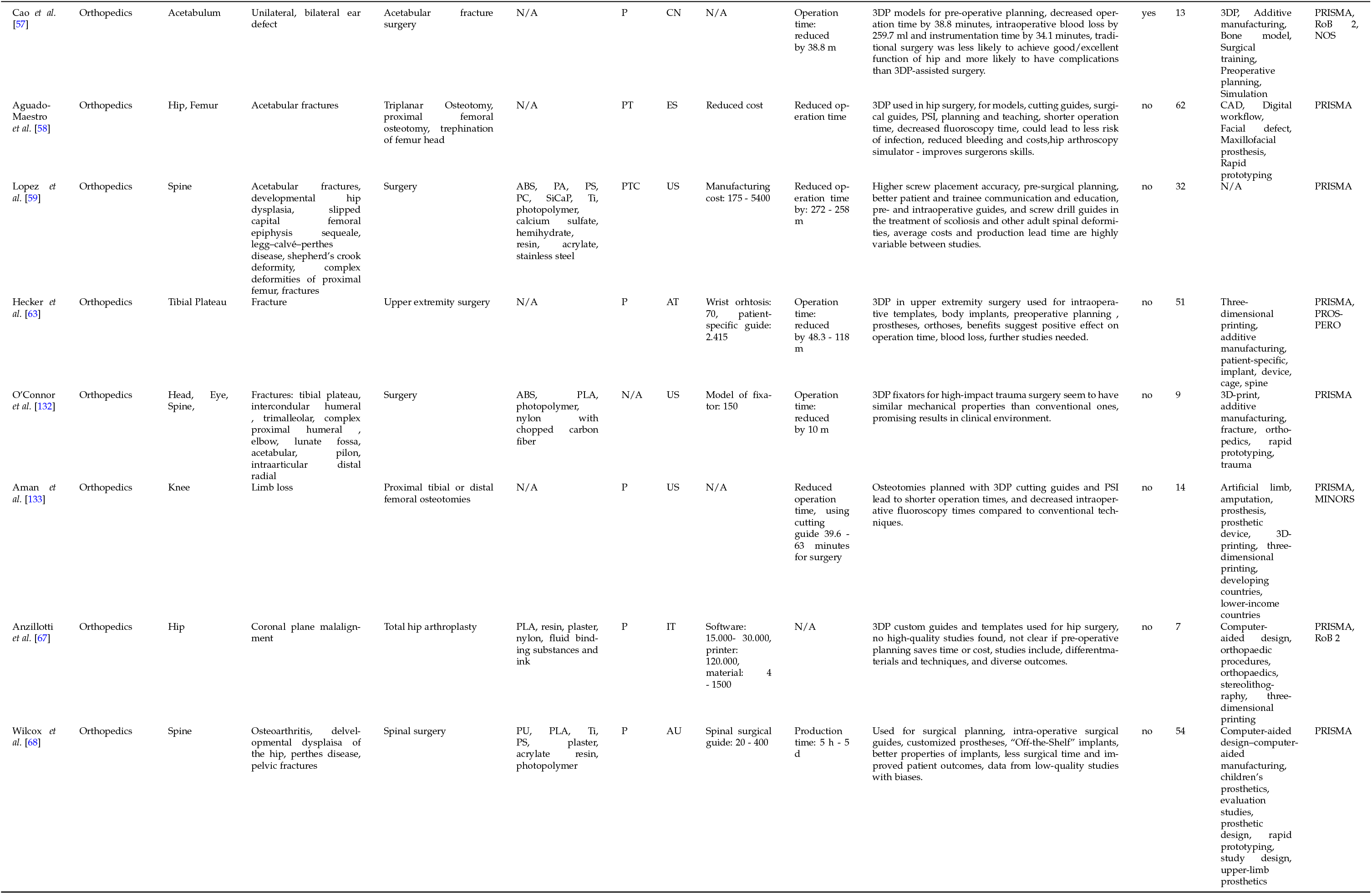
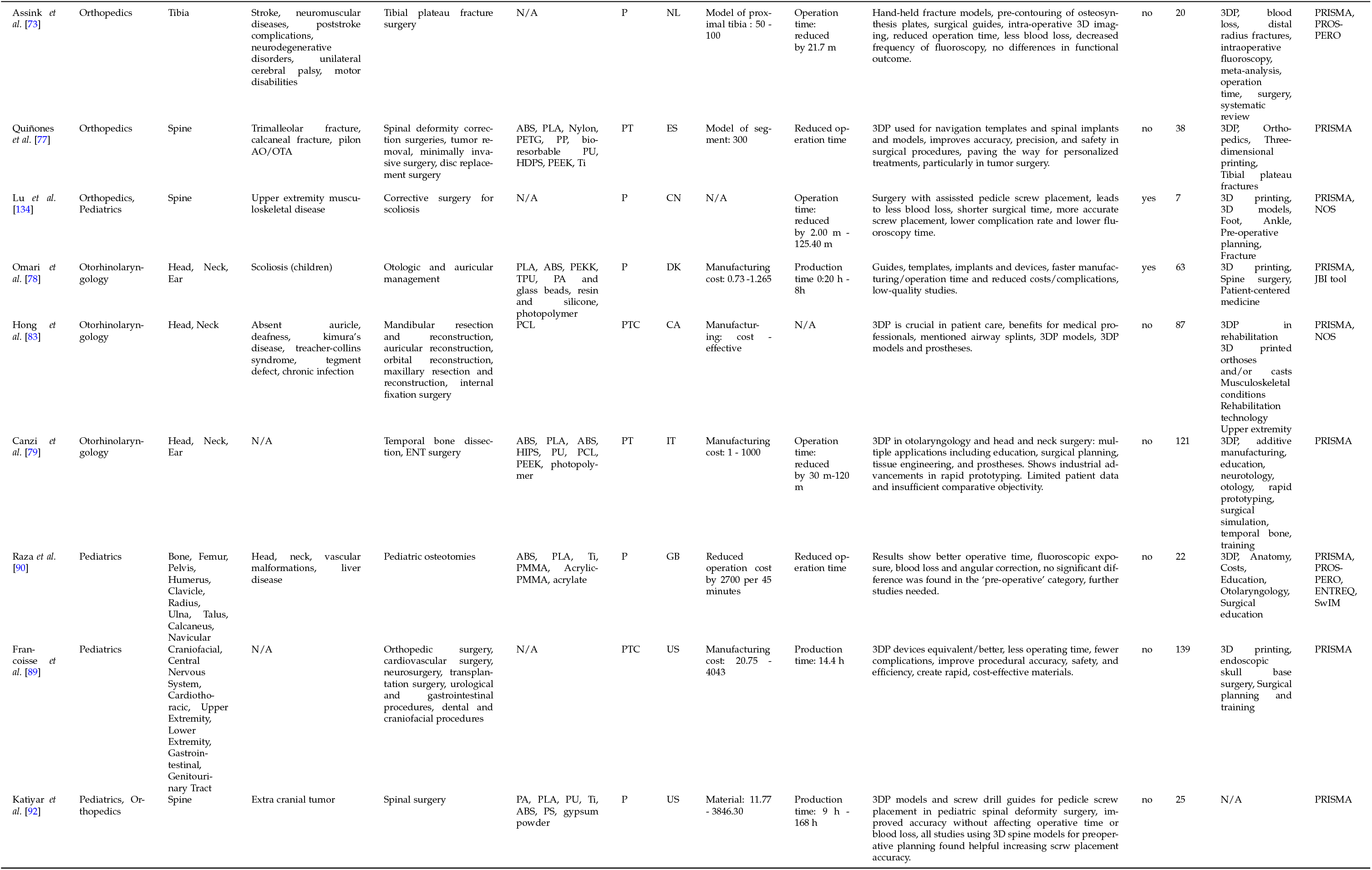
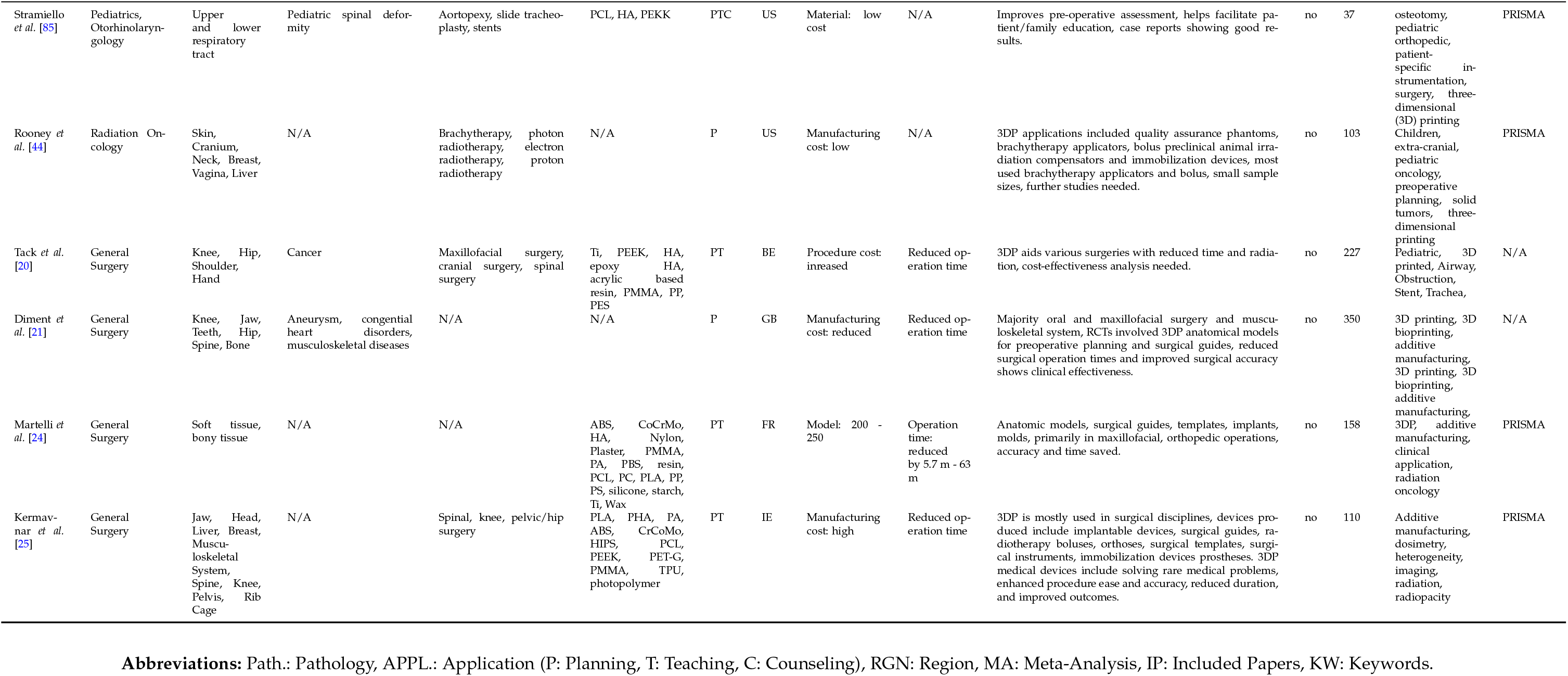
Systematic Reviews of Invasive and Non-permanent Applications.

**Table 4:**
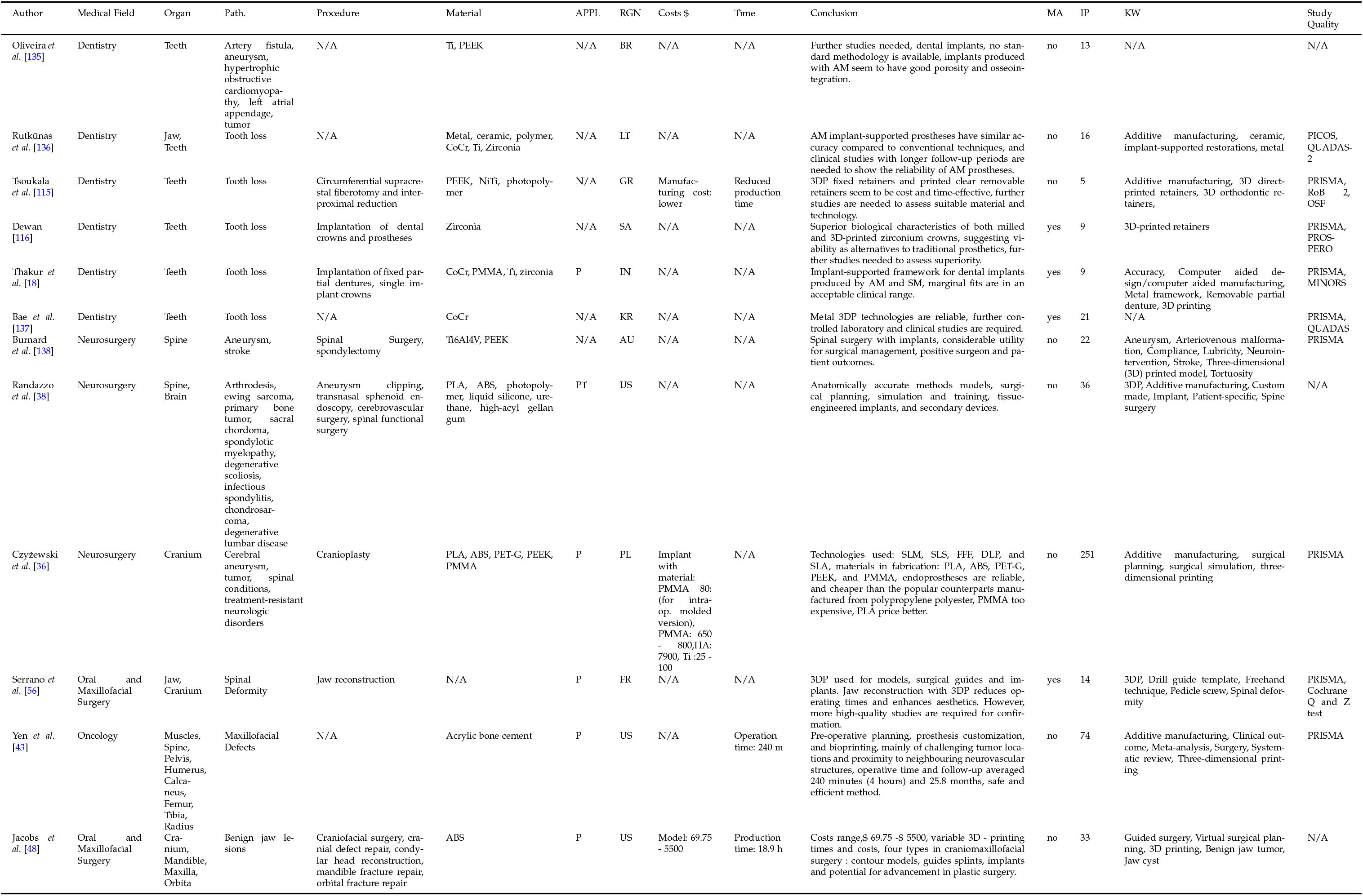
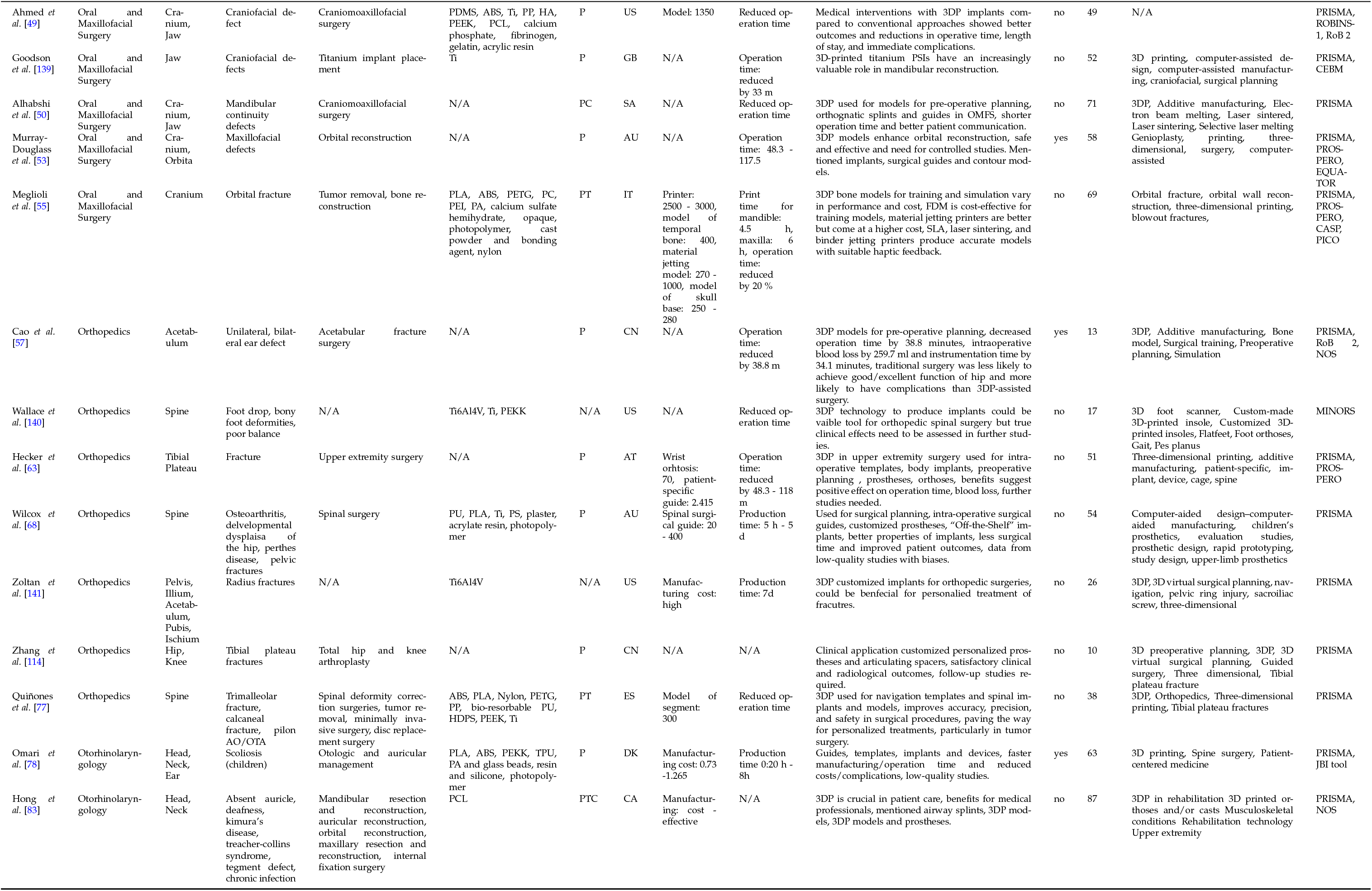
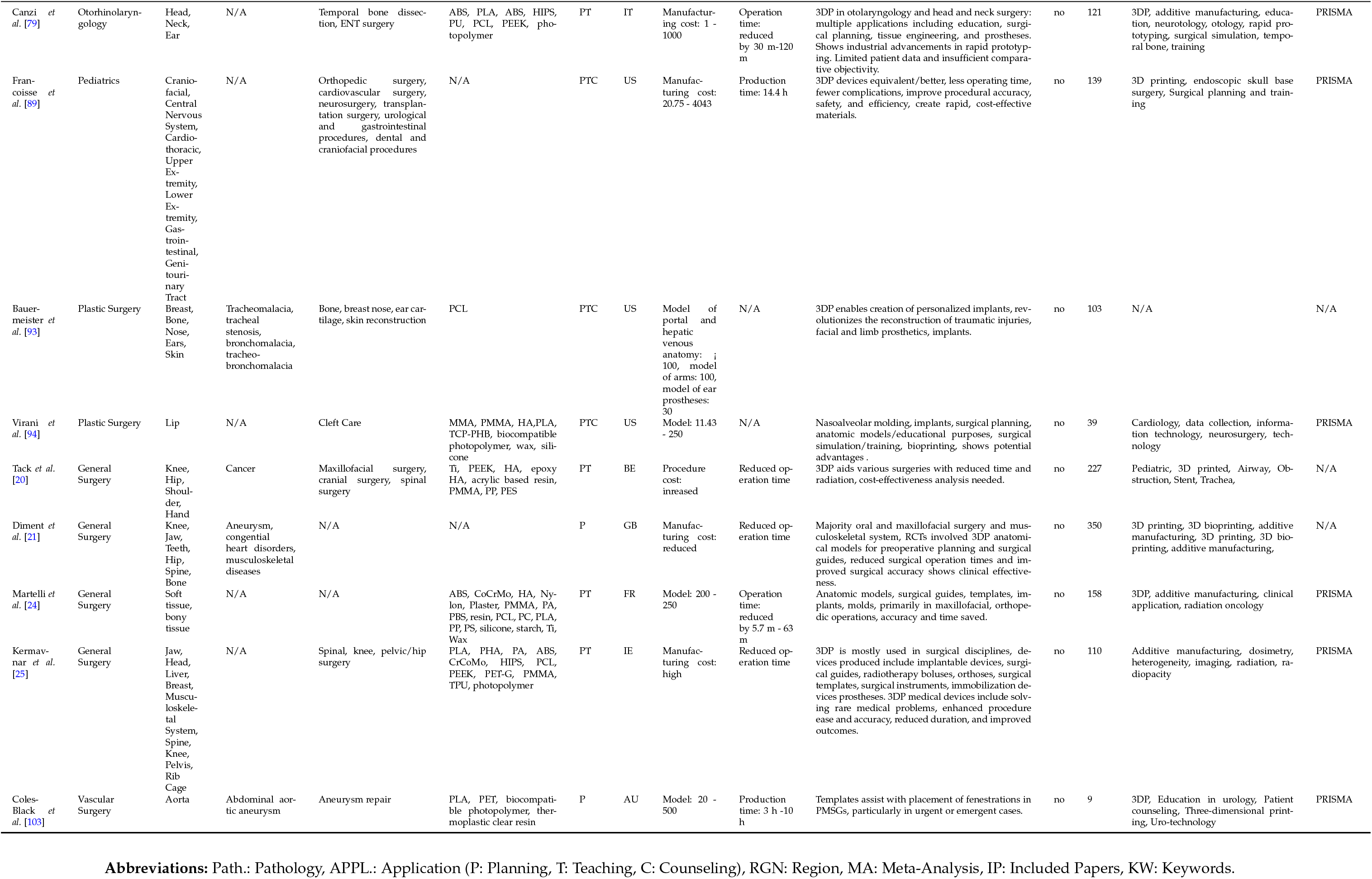
Systematic Reviews of Invasive and Permanent Applications.

**Figure 2.**
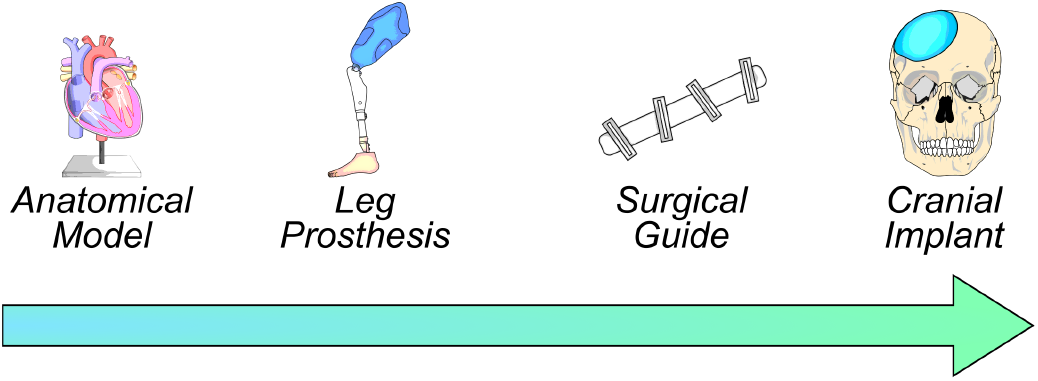
Degree of invasiveness. The heart was created by Eric Pierce and the prosthetic leg was created by Jean-Paul Gibert and color matched for the illustration. License CC-BY 4.0.

### 2.3 Reviews in Medical 3D Printing

#### 2.3.1 Non-Invasive Applications

In Table 1, reviews studying non-invasive methods are listed (n = 106). Non-invasive devices are external applications, like 3DP models. The medical fields included are anesthesia [1], cardiology [2–13], coloproctology [14], dentistry [15–18], general surgery [19–25], health care engineering [26, 27], hepatology [28], interventional radiology [29], medical education [30–34], neurosurgery [35–39], oncology [40–43] and radiation oncology [44–46]. Further medical fields included are oral and maxillofacial surgery [47–56], orthopedics [57– 77] and otorhinolaryngology [78–84]. The field of pediatrics [85–92] included sub-specialties such as, orthopedics [92], otorhinolaryngology [85, 86] and oncology [91]. Yang and colleagues [87] mentioned the following pediatric surgical sub-specialties: neurosurgery, cardiothoracic Surgery, plastic Surgery, cardiovascular surgery, urological surgery, orthopedics, ENT surgery, oncological surgery and vascular surgery. Further medical fields include plastic surgery [93, 94], traumatology [95–97], urology [98–102] and vascular surgery [103–106].

The affected organs or organ systems include the aorta [13, 103, 106], bladder [29], brain [35, 37, 39], cranium [30, 31, 34, 36, 44, 48–50, 53–55, 82, 84, 86, 91], ear [48, 78–81, 83,93, female internal genitals [46], heart [2–12, 87, 88, 104], jaw [17, 20, 21, 23, 25, 47–52, 55, 56], kidney [29, 41, 98, 100–102], lip [94], liver [19, 22, 28], musculoskeletal system [20, 21, 24, 31–33, 38, 43, 57–77, 89, 90, 92, 93, 95–97], prostate [98, 99], rectum and colon [14, 40, 42, 45], spine [38, 59, 68, 77, 92], teeth [15–18], thyroid gland [45], upper and lower respiratory tract [1, 45, 85] and vessels [39, 105].

The pathologies in this section include cardiovascular diseases [2–8, 10–12, 103–106] such as congenital heart disease, aneurysm and left atrial appendage. The second group includes cereberovascular diseases [35, 37, 39, 104], such as aneurysm, stroke and cranial defects. Further pathologies are musculoskeletal diseases [25, 36, 43, 47, 48, 50, 53–55,57–65, 67–77, 90–92, 95–97] which include different fracture types, such as pelvic fractures and tibial plateau fractures but also complex spinal deformities. Finally, diseases of the urogenital tract were found [29, 41, 46, 98–100, 102] such as renal disease.

Procedures include orthopedic surgery [25, 57–77, 95–97]. More precisely foot and ankle fracture fixation [76], tibial plateau fracture surgery [73, 74], open reduction and internal fixation surgery [75, 97], fracture surgery [57, 60–65, 69, 71, 72, 74, 95, 96], treatment of pelvic ring injuries [70], osteotomy [58, 66], total hip and knee arthroplasty [67]. Quinones and colleagues mention [77] complex enbloc resections of primary vertebral tumors, complex cran-iocervical junction surgeries, spinal deformity correction surgeries, fracture-dislocations of the thoracic and lumbar spine, minimally invasive surgery, disc replacement surgery. The next procedures included cardiovascular surgery [2–13, 103, 104, 106] are aneurysm repair [103, 104, 106], left atrial appendage closure [5, 10–12], transcatheter aortic valve replacement [8, 13] and intra-cardiac repair with surgical or catheter-based procedures [2]. Neurosurgical procedures [35–39, 104] include aneurysm clipping [35, 38, 104], cranioplasty [36] and open vascular and endovasular surgery [37]. Blohm and colleagues [35] add ventriculostomy and external ventricular drain placement, clinoidectomy, sagittal sinus repair, arteriovenous malformation repair, carotid artery dissection repair and hematoma repair. Another author mentions transnasal sphenoid endoscopy, cerebrovascular, spinal functional surgery [38]. Oral and maxillofacial surgical procedures and plastic surgery procedures [47, 48, 50–56,94] include orbital reconstruction surgery [48, 53, 54], jaw reconstruction [56] and cleft care [94]. Jacobs and colleagues [48] mention methods such as cranial defect repair, condylar head reconstruction and mandible fracture repair. In the field of otorhinolaryngology, procedures include temporal bone surgery [79–82]. De Souza and co-writers [81] mention other procedures laryngoplasty, middle ear surgery, FESS, subglottic stenosis treatment, labial cleft surgery and septoplasty. Other more specifically surgical procedures of the ear include ortic mastoidectomy, canaloplasty, labyrinthectomy, stapedotomy, tympanostomy and myringotomy, which Leung and colleagues [84] list. Procedures of the urogential tract involve prosteactomy [99], nephrectomy and laparoscopic partial nephrectomy [98, 102] as well as ureteral stent placement and kidney transplantation [98].

Within general and plastic surgery, gastroenterologic interventions include hepatic surgery and liver resection, living donor liver transplantation [19, 22, 28], as well as complete mesocolic excsision [14]. Procedures found in plastic surgery include breast, bone, nose, ear and skin reconstruction [93]. Radiation oncology procedures include brachytherapy, photon radiotherapy, electron radiotherapy and proton radio-therapy [44–46].

Materials can be categorised into 5 classes: polymers, metals, ceramics, organic materials and composite materials. In the class of polymers the following materials were found: ABS [1, 8, 37, 38, 45, 48, 61, 64, 79, 88, 90, 105], clear resin [103, 106], HIPS [79, 80, 84], latex resin [12], nylon [24, 28, 77, 105], PA [11, 28, 81, 92], PS [92], PET-G [25, 27, 77, 106], PCL [83, 84], PEEK [77, 92],PHA [25], photopolymer [5, 6, 12, 16, 25, 38, 79, 80, 103–106], PLA [4, 17, 25, 40, 55, 77, 80, 104, 105], PMMA [18, 49, 90, 94], PVS [15, 91], PU [4, 38, 92], PUR [105] and silicone [11, 13, 19, 38, 47, 49, 78, 80, 84]. Organic materials include biocompatible photopolymer [25, 94, 103, 106], gelatin [12, 49], high-acyl gellan gum [38], starch and cellulose powder [104] and wax [94]. Metals comprise CoCr alloy [18, 24, 26] and Titanium [18, 24–26, 49, 68, 77, 90, 92]. Composite material include TCP-PHB [94] and resins combining soft materials, rubber-like material and rigid materials [86]. The last material class of ceramics include HA [20, 85, 94], gypsum powder [81, 92], calcium sulfate hemihydrate powder [80, 81] and acrylic bone cement [43]. Table 1 shows 3DP for non-invasive applications. It was mostly used for pre-operative planning [2, 5, 10, 13, 15, 18, 19, 21, 22, 36, 39, 41, 43, 44, 46–48, 52–54, 56, 57, 60, 61, 63, 64, 67, 69–76, 78, 90, 92, 96, 97, 102, 103]. Other combinations found were pre-operative planning and teaching [3, 4, 7–9, 12, 14, 20, 24, 25, 28, 34, 35, 37, 38, 51, 55, 58, 62, 77, 79, 84, 86, 88, 104], pre-operative planning, teaching and patient counseling [6, 11, 42, 59, 83, 85, 89, 91, 93–95, 98, 99, 101, 105, 106], teaching[1, 29–33, 80–82], pre-opertive planning and patient counseling [40, 50, 65, 66, 87, 100] and patient counseling [68].

The top three countries, with most publications originated from, in alphabetical order, are Australia [1, 6, 7, 16, 22, 28, 45, 53, 68, 99, 101–104], China [9, 30, 41, 57, 60, 61, 65, 69, 72, 74, 75, 84, 96, 105, 107] and United States [29, 31, 35, 37–40, 43, 44, 48, 59, 94, 98, 108–110].

Overall, the costs had a broad range for a model from $ 0.6 - $5500 per model [1, 6, 23, 29, 37, 40, 48, 68, 84, 89, 91, 99–102, 105, 108, 110, 111] and the printer costs ranged from $500 - $200.000 [29, 42, 51, 105].The manufacturing cost range is $ 0.73 - $ 5400 [2, 62] and the material cots range is $ 1 - $ 7500 [35, 44, 57]. One study [1] made a cost analysis specific to anesthesia models. One author gave detailed information about the costs of 3DP in medicine [23].

The section time gathers information about reduced operation time, production time and specific print time for an 3DP object. The operation time was reduced by 0.72 - 258 minutes [42, 94]. The time to produce a 3D-printed model was 20 minutes to 9 days [28, 40, 88, 89, 91, 101, 102]. Morgan and colleagues [95] reported a reduction of the operation time by 19.85 %.

Studies reported positive effects on the operation time [3, 4, 20, 24, 41, 43, 56, 57, 60–62, 64, 65, 68–70, 72–75, 78, 89,90, 95, 96, 99, 112],blood loss [41, 60–62, 64, 65, 69, 70, 73, 75, 95, 96] and fluoroscopy dose [20, 61, 70, 90, 95, 96]. There were reports about less risk of infection [78] and better functional outcomes [24, 41, 52, 60–62, 65, 69, 74, 75, 89, 113]. Authors reported about teaching benefits [31, 80, 105]. A lot of authors point out that more high-quality studies/trials are needed [2, 9, 41, 53, 56, 75, 79, 88, 112, 114]. In addition, these authors conducted meta-analysis [3, 5, 7, 15, 30, 31, 41, 51, 53, 56, 57, 60, 61, 64, 65, 69, 72, 74, 75, 78, 81, 84, 91, 95, 96, 113]. The authors included between 4 to 350 articles.

These authors followed the PRISMA guidelines [1, 3, 5–7, 9, 13–19, 22–26, 28, 30–37, 39–45, 47, 49–79, 81–92, 94–106], were registered [4, 14, 16, 40, 47, 51, 53–55, 60, 63, 70, 72, 73, 84, 88, 90, 91, 95, 98, 99] and did a bias assessment [4, 5, 7, 14–18, 23, 29–31, 33, 39–41, 49, 51, 53–57, 60–62, 65, 67, 69, 71, 72, 74–76, 78, 82–84, 86, 90, 96, 97, 99, 100].

#### 2.3.2 On-body Applications

In Table 2, reviews are listed about on-body devices or applications (n = 26). On-body devices refer to applications that are in direct contact with the skin, such as prostheses. They include the following medical fields dentistry [18, 115–117], health care engineering [26, 27], general surgery [24, 25], oncology [42], oral and maxillofacial surgery [48, 50, 118], orthopedics [63, 74, 119–125], otorhinolaryngology [78, 83], pediatrics [89], plastic surgery [93] and radiation oncology [44]. The organs or organ systems in this section include the cranium [44, 48, 78], musculoskeletal system [24, 44, 63, 74, 93, 119–125], rectum [42], teeth [18, 115–117] and trachea [124]. In addition Francoisse and colleagues [89] mention organ systems such as the craniofacial region, central nervous system, cardio-thoracic system, musculoskeletal system and gastrointestinal and genitourinary tract.

The pathologies in this section include cancer [25, 42, 44, 83], Covid-19 [26, 27], cranio-maxillofacial defects [25, 48, 50, 118], foot abnormalities such as flat feet [120, 121], limb loss [122, 123], neurological disorders [124], tooth loss [18, 115–117], various fracture types such as forearm fractures or tibial plateau fractures [63, 74, 119, 125] and different types of ear disorders [78].

Procedures include dental procedures such as implantation and production of dental crowns or prostheses [18, 116, 117], production of personal protective equipment (PPE) such as face shields [26, 27] and surgical procedures such as total mesorectal excision, craniofacial surgery, upper extremity surgery, amputation, otologic and auricular management, cardiovascular surgery, neurosurgery, transplantation surgery, urological and gastrointestinal procedures, orthopedic surgery [25, 42, 44, 48, 50, 63, 78, 83, 89, 93, 118, 122–124].

Material classes and associated materials include metals such as CoCr [18, 26, 117], NiTi [115] or Ti [18, 26]. Ceramics include zirkonia [116]. Within the material class of polymers ABS [25, 48, 118, 124], EVA [120], PLA [25, 27, 42, 78, 83, 119, 121, 122, 125], PA [78], PC [24, 121], PCL [24, 83], PEKK [78, 124], PET-G [25, 27, 121], PHA [25], photopolymer [18, 26, 27, 42, 78, 115], PU [121] and TPE [124] were found.

Combined materials inlude PC-ABS, [42] glass beads, resind and silicone [78]. Lastly organic materials such as starch[24, 125] and wax [24] were identified.

Authors used 3DP as tools for pre-operative planning [18, 44, 48, 63, 74, 78, 118], pre-operative planning and teaching combined [24, 25], planning, teaching and counseling combined [42, 83, 89, 93] and planning and counseling [50]. Countries with the most publications (n = 5) in this section originated from the United States [44, 48, 89, 93, 125]. The cost per model was 69.75 $ - 5500 $ [48], the material cost 10 $ - 980 $ [122] and the production cost 0.73 $ - 4043 $ [78, 89]. The production time had a wide range from 0:20 h - 5 d [78, 125] and concerning the operation time the reduction was from 5.7 m - 63 m in the OR [24]. It was reported that 3DP costs less than conventional methods and that a more personalized fit for the prostheses was possible with this technology [122]. These authors conducted an additional meta-analysis [18, 74, 78, 116, 117] The included papers ranged from 5 - 158. Concerning the study quality, the following authors followed the PRISMA guidelines [18, 24–26, 42, 44, 50, 63, 74, 78, 83, 89, 115–118, 120–125], did a bias assessment [18, 25, 74, 78, 83, 115, 117, 120, 121, 125] and were registered [63, 116, 125].

#### 2.3.3 Invasive and Non-permanent Applications

Table 3 is sorted according to the medical field (n = 42). The medical fields are dentistry [126, 127], general surgery [20, 21, 24, 25] health care engineering [26], neurosurgery [38, 128–131], oncology [42, 43], oral and maxillofacial surgery [47–50, 52, 53, 55, 56], orthopedics [57–59, 63, 67, 68, 73, 77, 132–134], otorhinolaryngology [78, 79, 83], pediatrics [85, 89, 90, 92] and radiation oncology [44, 46].

The affected organs or body parts are the brain [38], cardiothoracic system [89], central nervous system [89], cranium [25, 48–50, 52, 53, 55, 56, 78, 83, 132], ear [78, 79, 132], eye [132], female internal genitals [44, 46], gastrointestinal system [89], genitourinary tract [89], jaw [25, 47–50, 56, 126], liver [25, 44], musculoskeletal system [20, 21, 24, 25, 43, 57, 58, 63, 67, 73, 90, 133], neck [44, 78, 83, 132], rectum [42], soft tissue [24, 25, 44], spine [25, 38, 59, 68, 77, 92, 128–131, 134], teeth [21, 126, 127] and upper and lower respiratory tract [85].

The pathologies included cancer [21, 25, 42–44, 46, 55, 77, 83, 130], COVID-19 [26], craniofacial defects [47, 48, 53, 56], different fracture types [57, 58, 63, 67, 73, 132], ear and hearing disorders [78], musculoskeletal malformations [21, 67, 68, 133], neurologic disorders [21, 25, 38], respiratory disorders [21, 79, 85], spinal disorders [59, 68, 92, 128, 129, 134] and tooth loss [126, 127].

Procedures include brachytherapy [44, 46], cereberovascular surgery [38], colorectal surgery techniques, such as transanal total mesorectal excision, total mesorectal excision, laparoscopic resection [42], craniomoaxillofacial surgery [20, 48– 50, 52, 53, 56, 79, 83], guides for dental implants [126], orthopedic procedures on the musculoskeletal system [25, 47, 55, 57, 58, 63, 67, 73, 133], orthopedic surgery, cardiovascular surgery, neurosurgery, transplantation surgery, urological and gastrointestinal procedures, dental and craniofacial procedures [89, 90], otologic and auricular management surgery [78], production of medical equipment [26], spinal surgery [25, 38, 68, 77, 92, 128, 129, 131, 134], thoracic airway reconstruction techniques, such as aortopexy or slide tracheoplasty[85] and tooth transplantation [127].

In the material class of polymers ABS [38, 46, 48, 52, 55, 59, 77, 79, 90, 130, 132], HDPS [77], HIPS [25], liquid resin [26], nylon [26, 55], PA [26, 42, 47, 67, 92, 130], PC [42, 46], PCL [78, 83, 85], PDMS [49], PEEK [49, 73, 77, 92], PEI [26, 46, 55], PEKK [78, 85], PETG [25, 55, 77], PHA [25], photopolymer [24, 26, 38, 42, 47, 59, 78, 132], PLA [26, 38, 42, 46, 55, 67, 77, 78],PMMA [20, 25, 90], PPSF [46], PS [68, 92],PU [52, 68], silicone [38, 47, 52] and TPU [25] were found. In the material class of ceramics acrylic bone cement [43], HA [20, 85] and plaster [67, 68] were found.

The material class of metals includes CrCoMo [24, 25], metal [26, 126], SiCaP [59], stainless steel [59] and Ti [20, 26, 49, 59, 90]. The material class of organic materials includes bio-resorbable PU [77], calcium sulfate hemihydrate [55, 59], cast powder and bonding agent [55] and wax [24]. Lastly composite materials include PC-ABS [42].

Concerning the applications, 3DP is used for only planning [21, 43, 44, 46–49, 52, 53, 56, 57, 63, 67, 68, 73, 78, 90, 92, 126, 127, 130, 133, 134], planning and teaching together [20, 24, 25, 38, 55, 58, 77, 79], planning and counseling [50] and planning, teaching and counseling together [42, 59, 83, 85, 89].

The most reviews found in this section are from the United States [38, 43, 44, 46, 48, 49, 59, 85, 89, 92, 130, 132, 133].

The corresponding costs for the different categories are listed as follows: material 11.77 $ - 3846.30 $ [92], model cost: 20 $ - 5500 $ [48, 130] software: 15.000 $ - 30.000 $, printer: 120.000 $ [67] and manufacturing cost: 0.73 $ - 4043 $ [78, 89].

Regarding time, a reduction in operating time of 10 minutes to 258 minutes was reported [59, 132] and a reported production time range of 5 hours to 5 days [68]. These studies conducted an additional Meta-Analysis [53, 56, 57, 78, 128–131, 134]. The included papers by the authors in this section ranged from 6 - 350.

Regarding the study quality, these authors followed the PRISMA guidelines [24–26, 42–44, 47, 49, 50, 52, 53, 55–59, 63, 67, 68, 73, 77–79, 83, 85, 89, 90, 92, 126–134]. Conducted a bias assessment [49, 53, 55–57, 67, 78, 83, 90, 126–129, 131, 133, 134] and were registered [47, 53, 55, 63, 73, 90, 126]. The following authors conducted an additional Meta-Analysis Murray-Douglass *et al*. [53], Serrano *et al*. [56], Cao *et al*. [57], Omari *et al*. [78], Azimi *et al*. [128], Yu *et al*. [129], Wallace *et al*. [130], Liang *et al*. [131], and Lu *et al*. [134]

#### 2.3.4 Invasive and Permanent Applications

In Table 4 invasive permanent applications are listed (n = 35). The medical fields are dentistry [18, 115, 116, 135–137], general surgery [20, 21, 24, 25], neurosurgery [36, 38, 138], oncology [43], oral and maxillofacial surgery [48–50, 53, 55, 56, 139], orthopedics [57, 63, 68, 77, 114, 140, 141], otorhinolaryngology [78, 79, 83], pediatrics [89], plastic surgery [93, 94] and vascular surgery [103].

The affected organs include the teeth [18, 115, 116, 135–137], cranium [25, 36, 48–50, 53, 55, 56, 78, 79, 83], jaw [25, 48–50, 56, 94, 136, 139], ear [78, 79, 83], spine [38, 43, 68, 77, 138,140] and different parts of the musculoskeletal system [20, 21, 24, 25, 43, 89, 141].

Pathologies found in this section, in alphabetical order are aneurysm [20, 38, 103], cancer [25, 38, 43, 55, 77, 83, 130, 138,141], craniofacial defects [25, 36, 48–50, 53, 56, 139], diseases of different organ systems, including nervous, circulatory, respiratory, gastrointestinal, skin and muscular-skeletal [20, 21, 25], musculoskeletal diseases [20, 57, 63, 135], otologic diseases such as deafness or kimura’s disease [78, 79], spinal disorders [25, 38, 68, 77, 79, 138, 141] and tooth loss [18, 115, 116, 136, 137].

Procedures in this section found are aneurysm repair [38, 103], cranio-maxillofacial surgery [20, 36, 48–50, 53, 55, 56, 79, 94, 115, 139] such as mandibular resection or reconstruction [83]. Further procedures are ENT surgery [78], implantation of dental crowns and prostheses [18, 116], orthopedic surgery [57, 63] such as total hip and knee arthroplasty [114]. In pediatrics, orthopedic surgery, cardiovascular surgery, neurosurgery, transplantation surgery, urological and gastrointestinal procedures, and dental and craniofacial procedures were found [89]. Further procedures include plastic reconstruction surgery on the breast or nose, ear cartilage and skin [93]. The last procedure found in this section includes spinal surgery [25, 68, 77, 138].

In terms of materials, they are listed by class: polymers, ceramics, metals, organic materials and composite materials. Polymers found are ABS [24, 48, 77, 79], HDPS [77], MMA [94], Nylon [24, 55, 78], PA [24, 25, 78], PBS [24], PC [20, 24, 49], PCL [24, 25, 78, 83, 93], PEEK [18, 20, 24, 25, 77, 79, 138], PEKK [78, 115, 140], PETG [24, 55, 77], photopolymer [20, 38, 55, 78, 94, 103], PLA [24, 25, 36, 55, 77, 103], PMMA [18, 24, 103], PP [20, 24, 78], PS [24, 68], resin [24, 49, 55, 78], silicone [24, 78, 94], TPU [25, 78] and zirconia [18, 116, 136]. In the material class of ceramic materials calcium phosphate [49], cast powder and bonding agent [55], HA [20, 24, 94], opaque [55] and plaster [24, 68] were found. With the material class of metals CoCr [18, 136, 137], Ti [18, 20, 24, 135, 140, 141] and NiTi [115] were derived from the papers. Organic materials include starch [24], wax [24, 94], fibrinogen [49], gelatin [49], and bio-resorbable PU [77]. The only compound material found in this section was TCP-PHB [94]. 3DP was used for pre-operative planning [18, 21, 36, 43, 48, 49, 53, 56, 57, 63, 68, 78, 103, 114, 139], pre-operative planning and patient counseling [50], pre-operative planning and teaching [20, 24, 25, 38, 55, 77, 79] and planning, teaching and patient counseling combined [83, 89, 93, 94].

In this section Australia was found to be the leading country with publications from [53, 68, 103, 138]. The costs found for the following materials are PMMA: 80 $ - 800 $, HA: 7900 $, Ti:25 $ – 100 $ [36]. Concerning the manufacturing cost a range from 0.73 $ to 4043 $ was found[78, 89]. The model cost range from 11.43 $ - 5500 $ [48, 94].

Concerning time the operation time was reduced by [24, 79] 5.7-120 minutes and the manufacturing time was 0.20 h -5 days [68, 78]. The 3D-printed implants seem to have acceptable accuracy [57, 89] and using the technology leads to less surgical time [57, 68, 89].

These authors conducted an additional Meta-Analysis [18, 53, 56, 57, 78, 116, 137].

The range of papers included by the authors is between 9 and 350. Furthermore, most studies followed the PRISMA guidelines [18, 24, 25, 36, 43, 49, 50, 53, 55–57, 63, 68, 77–79, 83, 89, 94, 103, 114–116, 137–139, 141]. Additionally these authors conducted a Bias Assessment [18, 49, 53, 55–57, 63, 78, 83, 115, 136, 137, 139, 140] and were registered [53, 55, 63, 116, 137].

## 3 Results

### 3.1 Medical fields and applications

3DP is used in a wide range of medical fields (Figure 3), procedures (Figure 6), pathologies (Figure 7), organs (Figure 4) and applications (Figure 5). The majority of these are disciplines surrounding surgical areas. This is the first medical discipline mentioned in our tables. These include anaesthesia (n = 1), cardiology (n = 12), coloproctology (n = 1), dentistry (n = 12), general surgery (n = 7), health care engineering (n = 2), hepatology (n = 1), interventional radiology (n = 1), medical education (n = 5), neurosurgery (n = 10), oncology (n = 4), oral and maxillofacial surgery (n = 12), orthopedics (n = 34), otorhinolaryngology (n = 7), pediatrics (n = 8), plastic surgery (n = 2), radiation oncology (n = 3), traumatology (n = 3), urology (n = 5) and vascular surgery (n = 4).

**Figure 3.**
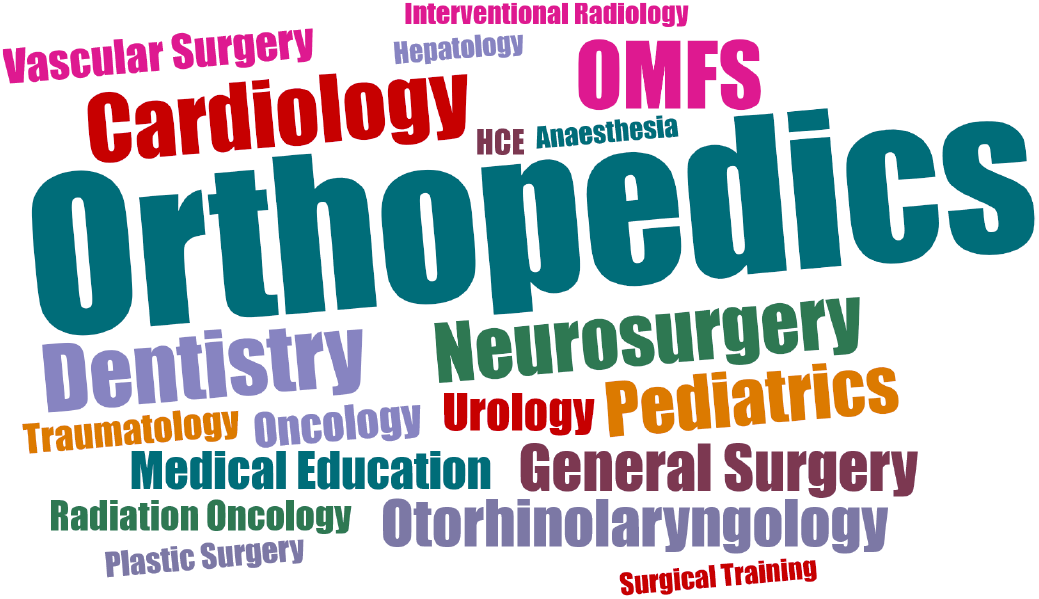
Medical fields breakdown.

**Figure 4.**
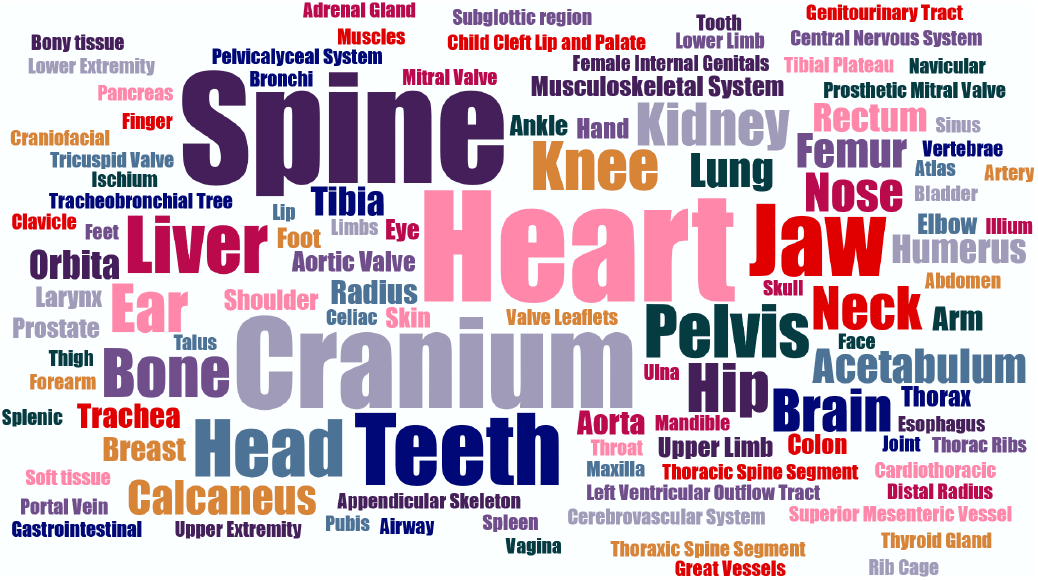
Organs breakdown.

**Figure 5.**
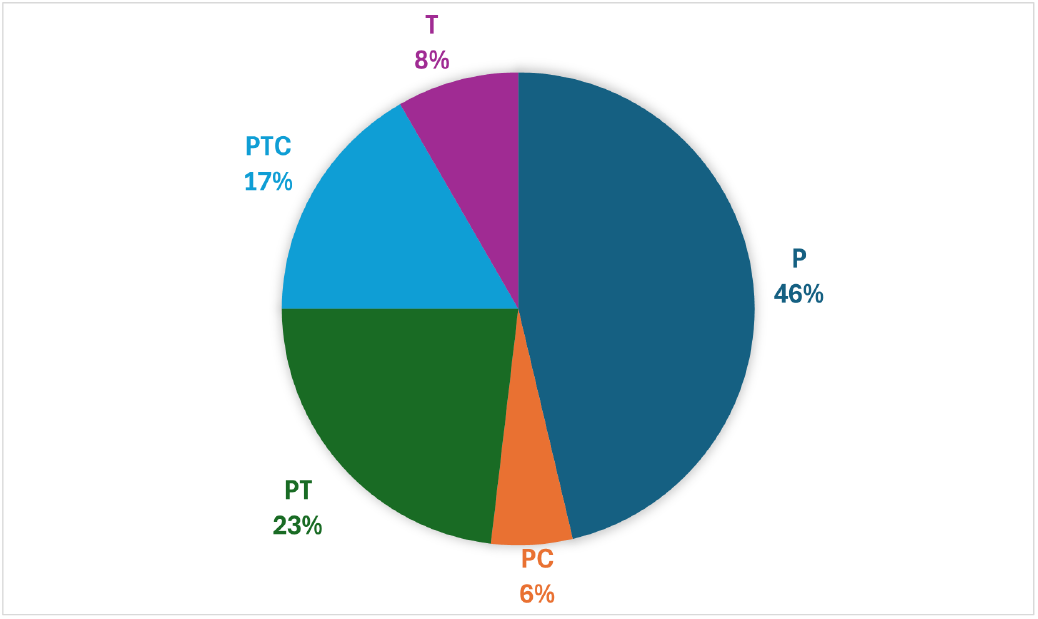
Distribution of applications including preoperative planning (P), teaching (T), or patient counseling (C).

**Figure 6.**
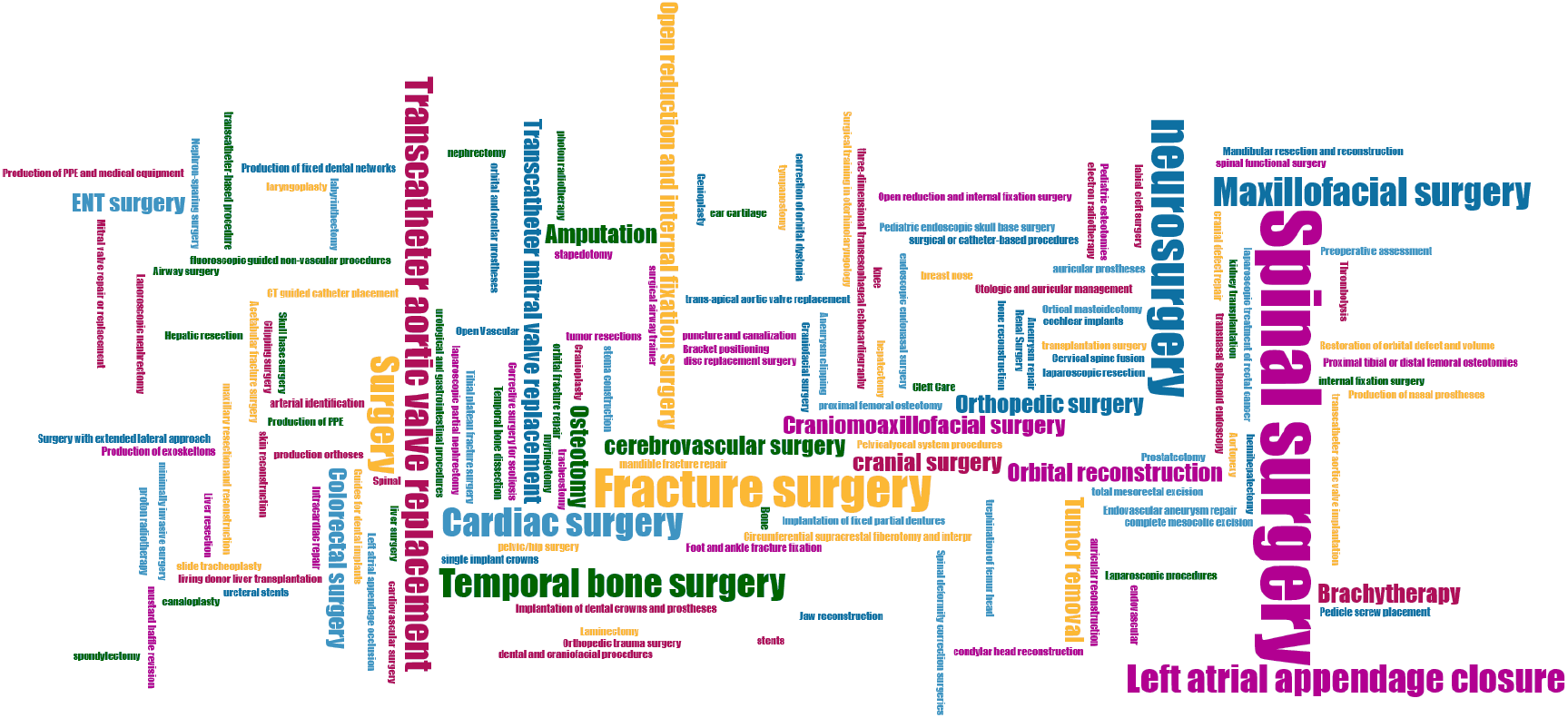
Procedures breakdown.

**Figure 7.**
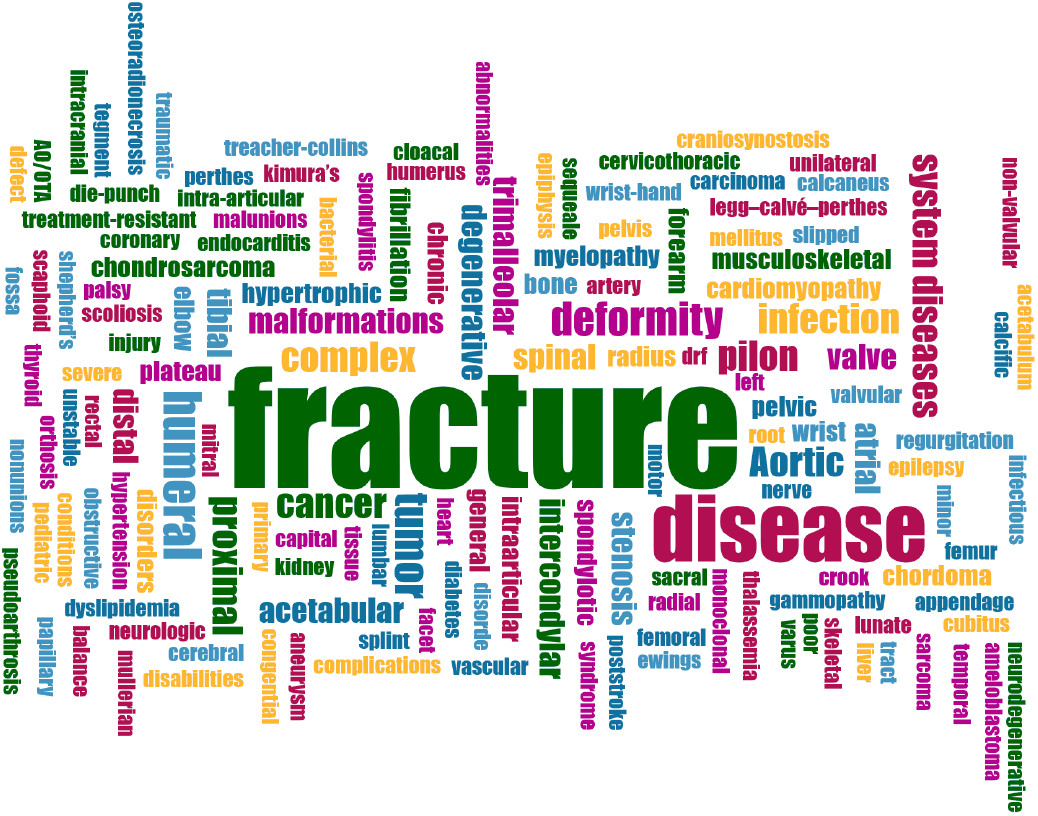
Pathologies breakdown.

#### 3.1.1 Orthopedics

The field with the most mentioned papers is the field of orthopedics (n = 34). 3DP can be used for models, pre-surgical planning, which can help the surgeon to prepare better for the surgery, because the 3D model makes it easier to understand the patient’s anatomy [68]. The 3DP technology is also used for prostheses. In this case, this field benefits from low material costs and better customization possibilities [114, 122]. Another application is individualized printed forearm braces used instead of traditional plaster casts, which might be more comfortable for the patient [119]. The last more invasive appliance for 3DP in this area are printed intra-operative guides such as screw drill guides or cutting guides as well as personalized implants [68, 141]. With the personalized guides, these surgeries can benefit from 3DP, in a way that for example the screws can be placed with more accuracy [59]. Other benefits in general are less blood loss during the surgery, less operation time and less fluoroscopy time [57, 76].

#### 3.1.2 Oral and Maxillofacial Surgery

Oral and maxillofacial surgery is the field with the second most amount of papers found in this meta-review. Main applications for 3D-printed models are intraoperative cutting/drilling guides and the use of patient specific implants (PSI) as part of the workflow in computer assisted surgery (CAS) [142]. Preoperative 3D-printed models with included pathology can serve as a preoperative planning tool or as an intra-operative reference. The pre-operative planned surgery is less invasive and seems to have acceptable accuracy [51, 52]. Other applications include in this medical specialty include printed surgical guides and individualized implants [48]. Otorhinolaryngology applies treatment to a similar area as Oral and Maxillofacial Surgery. Here, 3DP is also used for surgical guides, models, implants and personalized devices [79]. This leads to faster operation time and reduced costs and complications [79], and an overall improved surgical outcome.

#### 3.1.3 Dentistry

In dentistry 3DP is used to produce models, implants, such as crowns or prostheses and occlusal splints [126, 136].

#### 3.1.4 Cardiology

In cardiology, 3DP technology was only found to be used for models to pre-operatively plan the surgery [4]. With congenital heart disease, these models seem to be a valuable tool to help the surgeon better visualize the patient’s anatomy [2, 3]. The models seem to accurately replicate the anatomy [6]. It is also a helpful tool for teaching and patient counselling. There is a lack of studies in this field and more trials are needed to confirm the accuracy of these findings [9].

#### 3.1.5 Neurosurgery

In neurosurgery, 3DP is primarily used to create patient-specific models for pre-surgical planning [38]. These models also serve as valuable tools for medical training, allowing for the education of residents [37]. Additionally, 3D-printed models are utilized to produce surgical guides that assist surgeons during procedures, as well as implants for the field [130].

The use of 3D-printed models for pre-surgical planning has been shown to improve surgical times and patient outcomes. [129, 138]. For instance, in spinal surgery, 3D-printed surgical guides and screw placement guides are employed to achieve high screw placement accuracy, leading to reduced operation times and less blood loss [129].

#### 3.1.6 Pediatrics

Pediatrics is one of the areas where few reviews about 3DP were found. In this field, 3DP is used for pre-operative models, surgical guides and implants [89]. As children continue to grow, this has to be taken into account when planning operations involving implants. But also during surgery, 3DP can save time by pre-planning [89]. The accuracy of the models also seems to be good and inexpensive materials are used [89]. In the fields of anesthesia and interventional radiology and radiation oncology 3DP was used to produce models [29].

#### 3.1.7 Oncology

In oncology, 3DP is mainly used to produce 3DP models, which helps the surgeon prepare for the surgery. This technology is mostly used for the treatment of challenging tumor locations such as kidney and liver tumors, as well as the treatment of gastrointestinal cancer [28, 40, 41]. In special cases such, as the reconstruction of hemipelvis structures due to oncological diseases, a custom-made or 3D-printed endoprosthesis is used [143]. 3D-printed models have a high degree of accuracy and can be a very valuable tool in patient education, as well as preoperative planning, which could shorten operating times [41].

#### 3.1.8 Urology and Vascular Surgery

In the fields of urology and vascular surgery the technology is used to print 3D models, which are used for patient counseling, pre- and intra-operative surgical planning, as well as educational purposes [42, 43]. These fields are not very often mentioned and the use of implants or other 3D-printed devices is not reported yet.

#### 3.1.9 Traumatology

Traumatology treats similar parts of the body as orthopedics, but the injuries are evoked with higher impact forces.3DP is again a good tool to be used to print fracture models to help the surgeon understand the complex anatomy [96, 97]. When pre-planning a surgery with 3D models, it was reported that there was less fluoroscopy time, blood loss, and operation time [95, 97].

### 3.2 Grade of Invasiveness

The reviews were classified by the grade of invasiveness. This turns out to be applicable for most of the reviews, but in some cases (n = 41), there were overlaps as some of them not only mentioned devices within one grade of invasiveness, but multiple grades of invasiveness. Therefore, some reviews were mentioned multiple times.

#### 3.2.1 Non-invasive Applications

The largest area of 3DP applications in medical fields are non-invasive applications that do not involve physical contact with the patient. Here, the medical field of orthopedics is the leading one, followed by cardiology, oral and maxillo-facial surgery. In these fields, 3DP is mainly used for pre-operative planning with patient-specific models that help the surgeon gain a better understanding of the anatomy of the patient [50, 66]. But it is not possible to say which organ or pathology is most often treated with the help of 3DP.

#### 3.2.2 Invasive Non-permanent

In the field of invasive non-permanent devices, orthopedics and oral and maxillofacial surgery are the most mentioned fields. Non-invasive permanent devices are designed to remain within the body only for a short period, for example, during surgery. These include operational devices such as cutting guides, screw guides, or personalized surgical instruments [56, 58].

#### 3.2.3 Invasive Permanent

The medical fields of oral and maxillofacial surgery and orthopaedics are equally represented in the field of invasive permanent with the number of papers found (n=32) that are designed to remain in the body. These devices include personalised implants [53, 114, 140]. Again, positive effects on blood loss during surgery, time and outcome have been reported [49, 57, 63].

#### 3.2.4 On-body Applications

In terms of on-body applications, which are designed to come into contact with the skin, that include removable objects such as prostheses or occlusal splints, the medical speciality most frequently cited is orthopaedics. This is followed by dentistry and oral and maxillofacial surgery.In orthopaedics, the main use of 3DP at this level of invasiveness is for personalised prostheses or orthoses for patients with limb loss [122]. In oral and maxillofacial surgery, ear and nose prostheses are used [118].

### 3.3 Printing Techniques and Materials

Regarding the 3DP techniques and materials this topic is only briefly mentioned, as we focused more on the medical fields and applications. For completeness the following papers are included [27, 72, 144–149]. The materials used in bioprinting are alginate, gelatin and GelMA [144, 147]. The most used bioprinting techniques are pneumatic extrusion and piston [147]. But these techniques are in the early stages and few in vivo studies are reported [147]. For conventional 3DP printing, many materials are mentioned (Figure 8). The materials can be divided into polymers, ceramics and metals [145]. The most mentioned materials are ABS, titanium, and PCL [27, 149]. For 3DP technology, SLA and FFF seem to be the most used [146, 149]. But there are countless 3DP techniques and materials. It is difficult to get a clear result which material and 3DP method is the best. This study focuses on the use in different medical fields rather than the materials and technologies. As a result, this topic should be re-studied in an own contribution, to get a better overview.

**Figure 8.**
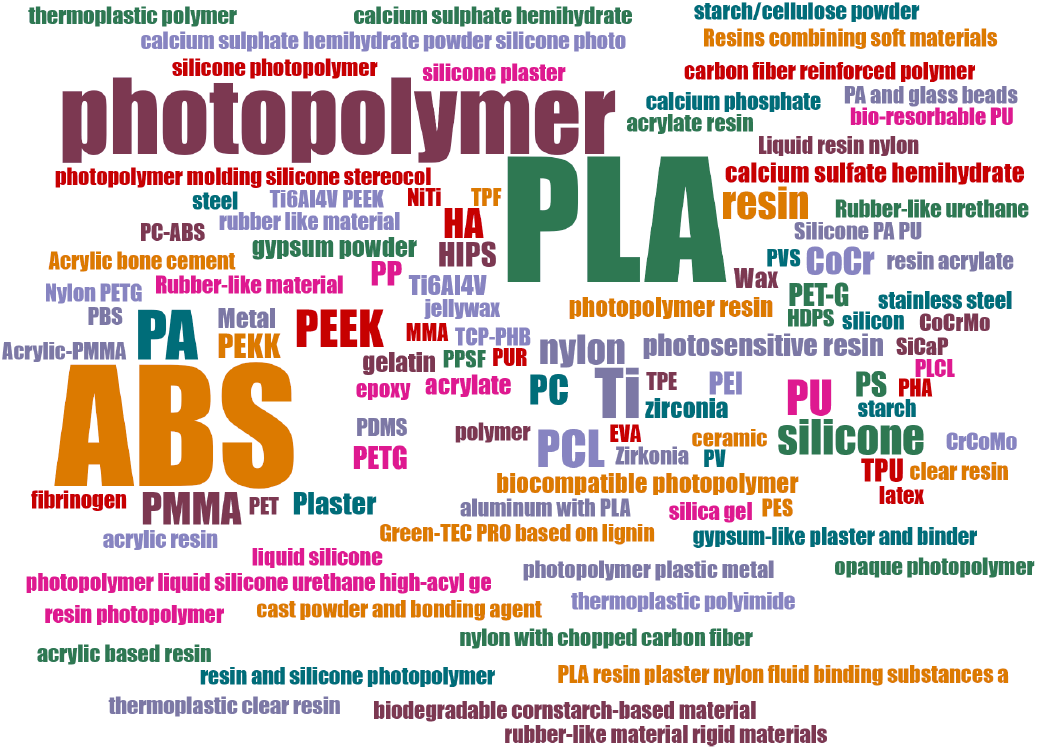
Printing materials breakdown.

### 3.4 Region

Abbreviations for countries have been chosen according to the PubMed ISO Country Codes for Selected Countries table [150]. The reviews of this meta-review were from a variety of countries, with the United States, China and Australia leading the way (Figure 9).

**Figure 9.**
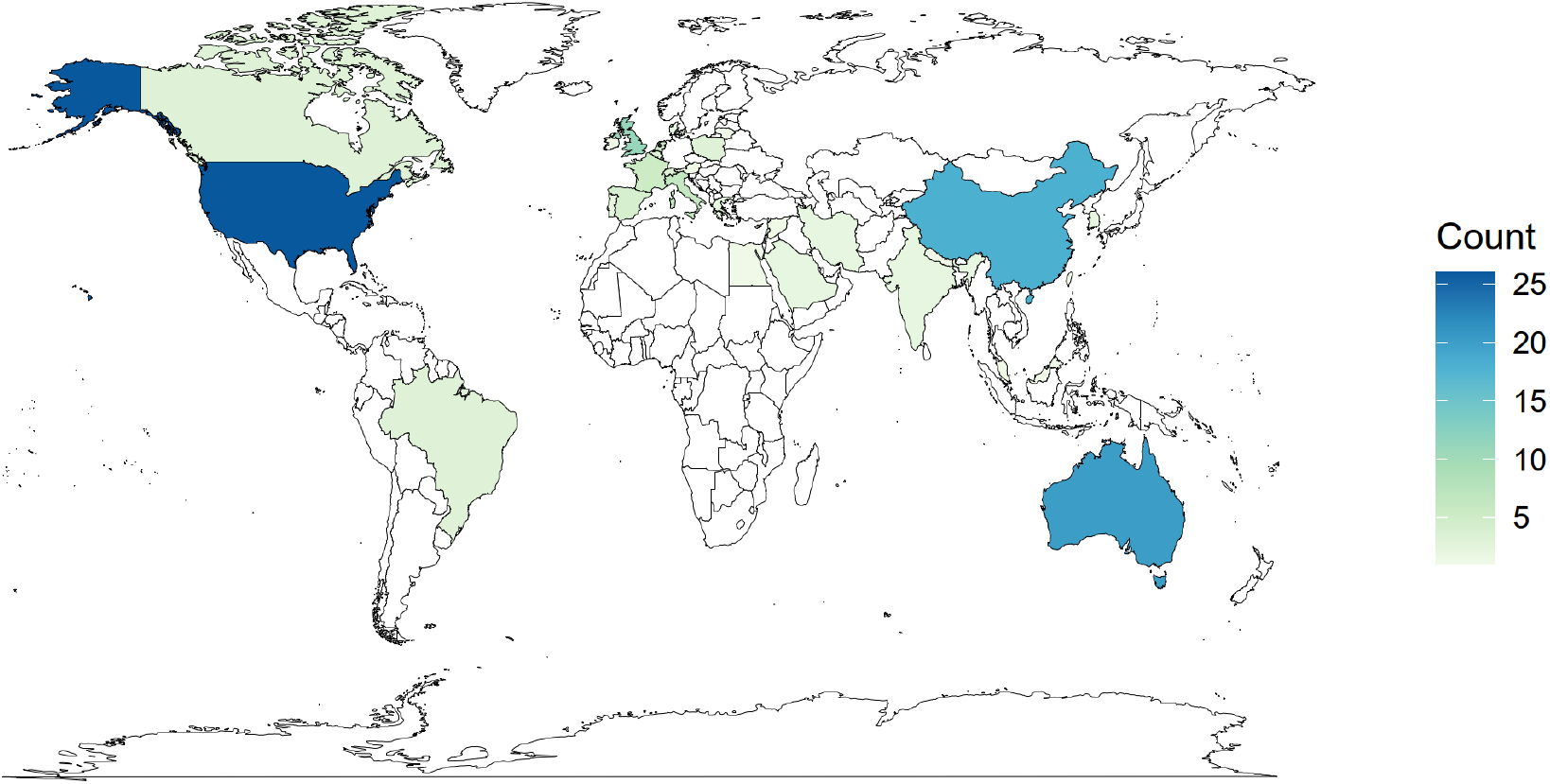
Origins of systematic reviews.

### 3.5 Keywords

The various systematic reviews were characterised by a large number of different keywords, in particular the terms “3D printing” and “additive manufacturing” (Figure 10).

**Figure 10.**
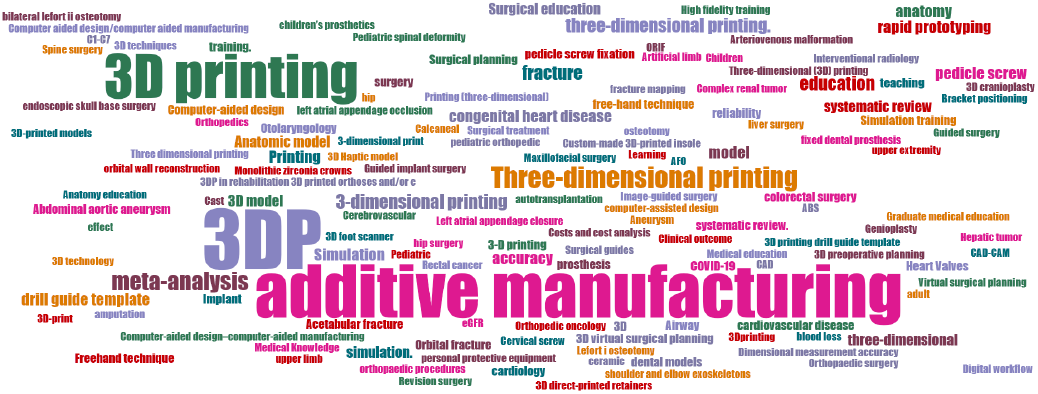
Keywords breakdown.

### 3.6 Study Quality

Concerning the study qualities the majority of authors followed the PRISMA guidelines only 18 did not follow them. Often the search criteria were not clear and they did not do a bias assessment either, so more high-quality studies and also randomized controlled trials (RCTs) need to be done to make the heterogeneous data more homogeneous (Figure 11).

**Figure 11.**
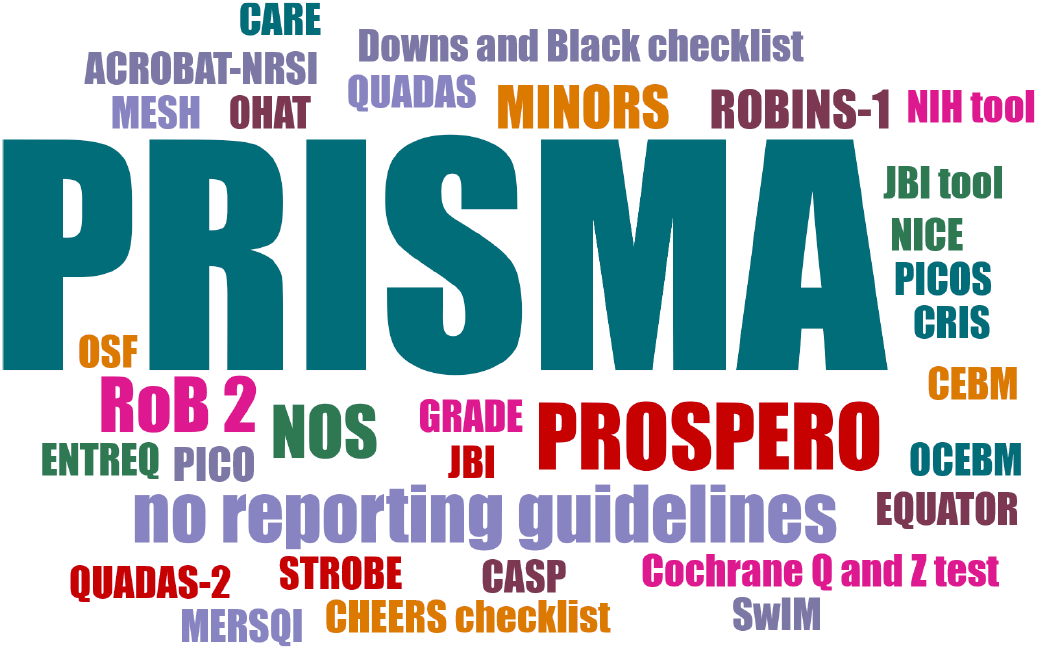
Quality indicators breakdown.

## 4 Discussion

The results of the study provide insights into which medical specialties use 3DP and to what degree of invasiveness. In addition, a rough overview of the materials and 3DP processes was given. The majority of medical disciplines using 3DP were centered around surgical fields. The medical specialties with the highest density of reviews found were orthopedics, cardiology, oral and maxillofacial surgery and dentistry. Very sensitive areas of the human body are often involved in these procedures. It is unclear, however, whether these fields are mentioned this often in the context of 3DP, because the technology is most prevalent in this area, or because it offers the most benefits. The classification of studies by degree of invasiveness is useful, because there are a number of different types of 3D-printed tools that can be accurately categorized by degree of invasiveness. We start with the non-invasive devices, which are shown in Table 1 and represent the largest group in our analysis. This grade of invasiveness is mostly represented by 3D-printed models. These models may be easier to produce and use, with fewer safety requirements, as these tools are not in physical contact with the patient [21]. The models are mainly used for pre-surgical planning. Concerning the accuracy, which is an important factor when planning a surgery with these devices, is to correctly represent the anatomy. Studies reported that the accuracy of these models was comparable or better than conventional methods [51]. This can help the surgeon to understand the patient’s anatomy or pathology better [128]. As a result, authors found that preparing a surgical intervention with 3D-printed models beforehand, can lead to less blood loss during surgery, a reduced surgical time and often a better surgical outcome, which has been shown in studies in various medical specialties [73]. Models can also be used for patient counseling, with a patient-specific model it is easier to explain surgical procedures to the patient [37]. Another use of 3D-printed models is for teaching students. Although these models seem to be cheaper, care should be taken to produce anatomically accurate models. No study was found that reported on the accuracy of teaching models. More attention should be paid on this topic. As technology progresses and the accuracy of these models is validated these 3D printed models could be used as an additional form of imaging technique in medicine. 3D-printed on-body devices such as prostheses or orthoses are represented in Table 2. Only 26 reviews were categorized here. Orthopedics is the dominant field in this type of invasiveness as this is the area where the need for prostheses is the greatest. Only one study compared these devices to conventional devices [121]. As a result of that more studies need to be conducted to assess the real mechanical properties, costs and wearability of these custom-made prostheses. Invasive devices can be categorized as invasive non-permanent devices (see Table 3) and invasive permanent devices (see Table 4). Here, only a minority of reviews could be classified in these categories. In invasive permanent devices, which are implants, the fields of neurosurgery and oral and maxillofacial surgery are the most mentioned ones. Customizing implants in these fields can be very useful for the treatment of patients and long-term outcomes. However, less is reported about the fabrication of these implants and how long it takes to produce an implant. Another issue is the sterilization process and the risk of infection when using 3D-printed implants. It is not clear yet which materials are the best that combine printability, sterilization and durability. Furthermore, none of the studies reported on the long-term effects of having 3D-printed implants in the body especially the ones made of different types of polymers. Further studies need to be conducted on what is the tolerability of these (polymer) implants’ risk of infection and the lifespan of 3D-printed custom-made implants. As a result, standardized safety regulations need to be assessed for these new tools. Invasive non-permanent devices are tools like cutting guides or screw guides that are designed to stay within the body but only during the period of the surgery and are then removed. Nothing was found about the sterilization process or the risk of infection in the literature, which has to be assessed in future studies. But literature suggests that spinal surgeries using screw guides have higher screw placement accuracy, which leads to a better surgical outcome [59]. After solving these problems and implementing safety regulations, these cutting guides could be used in standard surgical procedures in the future. In general, 3D-printed invasive devices are not yet used in standard medical practices as technology needs to be more sophisticated and more safety studies need to be done. It is difficult to make a statement about costs and time as the information in the literature is not standardized. Take the production of a model as an example. The materials might be cheaper but the work process is not fully described yet. We know that the data is derived from, for example, MRI or CT files and then a virtual model is produced or printed [138]. But the time it takes to produce the implant from acquiring images of the patient to the actual 3D-printed piece and the work of additional staff, such as the technician, has to be considered. This has also to be taken into account in terms of cost-effectiveness, as there are several steps involved in the printing process. In terms of time, sometimes the total time for the whole surgery using 3DP was given, and sometimes how much time was saved, but it was not exactly compared to surgery without using 3DP. Concerning materials and printing techniques there has to be a differentiation made between conventional 3DP with non-organic materials such as polymers and bioprinting with organic materials or even living cells. While bioprinting is still in its infancy, conventional 3DP is the more mature technology and is already being used in some medical settings.For conventional printing, materials like ceramics, polymers and metals are used [145]. Specifically, ABS, photopolymers and PLA are the most commonly mentioned materials. Most of these materials do not have antibacterial properties itself, the sterilisation process, as mentioned above, needs to be evaluated, but nothing substantial has been found in the literature. For the conventional 3DP technology, SLA and FFF seem to be the most used [149]. Multiple other techniques have been reported and the type of tool they are used for. However, we will not go into detail on these as this study focuses on the use of 3DP in different medical specialties. It has been shown that 3DP in general is used in various surgical fields. As the problems with the printing process, materials and costs are solved this technology could be promising for future medical practice.

### 4.1 Limitations

A limitation of our study is that our search was limited to the PubMed database. As a result, it is possible that some relevant studies may have been missed and are therefore not addressed in this meta-review.

## 5 Conclusion

In recent years, the scope of research and use of 3DP has continuously increased in the medical field. It offers a wide range of applications in pre-operative surgical planning, patient counseling and educational purposes. With its ability to produce patient-specific models, prostheses, surgical guides and implants, opening the door to personalized medicine. This can be seen in various medical fields, especially in surgical applications and medical education. The medical fields most frequently mentioned in the literature include orthopedics, oral and maxillofacial surgery, cardiology and neurosurgery. Non-bioprinting materials often include ABS, photopolymers and PLA. The literature suggests that surgical procedures benefit from reduced operating time, reduced blood loss, minimized fluoroscopy time and improved surgical outcome. In terms of time and cost, no consistent and accurate statement can be made due to the heterogeneity of the available data. Countries such as the United States, Australia and China have been at the forefront of these innovations. Future research and larger studies are needed to evaluate the clinical benefits of 3DP applications in medicine.

## Data Availability

All data produced in the present work are contained in the manuscript

## Acknowledgments

This work was supported by the REACT-EU project KITE (Plattform fü r KI-Translation Essen, EFRE-0801977, https://kite.ikim.nrw/), FWF enFaced 2.0 (KLI 1044, https://enfaced2.ikim.nrw/), AutoImplant (https://autoimplant.ikim.nrw/) and “NUM 2.0” (FKZ: 01KX2121). Behrus Puladi was funded by the Medical Faculty of RWTH Aachen University as part of the Clinician Scientist Program. Furthermore, we acknowledge the *Center for Virtual and Extended Reality in Medicine* (ZvRM, https://zvrm.ume.de/) of the University Hospital Essen.

